# Recipient APOL1 risk alleles associate with death-censored renal allograft survival and rejection episodes

**DOI:** 10.1101/2021.05.07.21256570

**Authors:** Zhongyang Zhang, Zeguo Sun, Qisheng Lin, Khadija Banu, Kinsuk Chauhan, Marina Planoutene, Chengguo Wei, Fadi Salem, Zhengzi Yi, Jia Fu, Ruijie Liu, Haoxiang Cheng, Ke Hao, Philip O’Connell, Shuta Ishibe, Weijia Zhang, Steven G. Coca, Ian W. Gibson, Robert B. Colvin, John Cijiang He, Peter S. Heeger, Barbara Murphy, Madhav C. Menon

**Affiliations:** Department of Genetics and Genomic Sciences, Icahn School of Medicine at Mount Sinai, New York, NY 10029, USA; Icahn Institute for Data Science and Genomic Technology, Icahn School of Medicine at Mount Sinai, New York, NY 10029, USA; Division of Nephrology, Department of Medicine, Icahn School of Medicine at Mount Sinai, New York, NY 10029, USA; Department of Pathology, Icahn School of Medicine at Mount Sinai, New York, NY 10029, USA; Westmead Millennium Institute, University of Sydney, NSW, Australia; Department of Medicine, Yale University school of Medicine, New Haven, CT 06519, USA; Department of Pathology, University of Manitoba, Winnipeg, Manitoba, Canada R3E 3P5; Department of Pathology, Massachusetts General Hospital, Harvard Medical School, Boston, MA 02114, USA

**Keywords:** APOL1, G1/G2 risk alleles, kidney transplantation, TCMR, allograft loss

## Abstract

Apolipoprotein L1 (APOL1) risk alleles in donor kidneys associate with graft loss but whether recipient risk allele expression impacts kidney transplant outcomes is unclear. To test whether recipient APOL1 allelic variants independently correlate with transplant outcomes, we analyzed genome-wide SNP genotyping data of donors and recipients from two kidney transplant cohorts, Genomics of Chronic Allograft Rejection (GOCAR) and Clinical Trials in Organ Transplantation 1/17 (CTOT1/17). We estimated genetic ancestry (quantified as proportion of African ancestry or pAFR) by ADMIXTURE and correlated APOL1 genotypes and pAFR with outcomes. In the GOCAR discovery set, we observed that the number of recipient APOL1 G1/G2 alleles (R-nAPOL1) associated with increased risk of death-censored allograft loss (DCAL), independent of genetic ancestry (HR = 2.14; P = 0.006), and within the subgroup of African American and Hispanic (AA/H) recipients (HR = 2.36; P = 0.003). R-nAPOL1 also associated with increased risk of any T cell-mediated rejection (TCMR) event. Analysis of the CTOT cohort validated these associations. *Ex vivo* studies of peripheral blood mononuclear cells revealed unanticipated high APOL1 expression in activated CD4^+^/CD8^+^ T cells and natural killer cells. We detected enriched immune response gene pathways in G1/G2 allele carriers vs. non-carriers among patients on the kidney waitlist and healthy controls. Together our findings highlight a previously unrecognized contribution of recipient APOL1 risk alleles to renal allograft outcomes. This immunomodulatory role has broader implications for immune mediated injury to native kidneys.

## INTRODUCTION

Patients with African ancestry sustain a significantly higher risk of non-Mendelian focal segmental glomerulosclerosis (FSGS) as well as end stage kidney disease (ESKD) (reviewed in literature (1)). Seminal work identified the risk genotypes of Apolipoprotein L1 (APOL1) when present as two copies of either or both G1 and G2 alleles,(2) explained the increased risk of FSGS and ESRD observed in African Americans (AAs).(3, 4) Mechanistic data have since focused on the role of APOL1 risk alleles in kidney epithelial cells including gain-of-function roles in FSGS(5) and pre-eclampsia,(6) and loss-of-function role in parietal cell biology.(7)

In renal transplantation, two copies of the APOL1 risk alleles when present in donor have associated with death-censored allograft loss (DCAL).(8-10) While donor African ancestry is incorporated into the kidney donor risk index,(11) giving weight to donors carrying two copies of APOL1 risk allele vs. all others, improved the prediction of DCAL.(12) Few mechanistic data suggest the development of FSGS in APOL1 risk genotype-carrying allografts.(13, 14) On the other hand, a single retrospective study in kidney transplant recipients reported that recipient carriage of APOL1 risk alleles was not associated with DCAL.(15) These data have since led to emphasis on the role of APOL1 expression in renal cells and outcomes. The role of APOL1 risk alleles in non-renal tissues including immune cells has not been specifically examined. A universal mechanism linking APOL1 risk alleles to allograft outcomes has not yet emerged from literature and a nationwide prospective study is currently underway.(16)

Previous studies by several groups show associations among self-declared AA ethnicity and increased rejection episodes and/or DCAL.(17-21) We recently reported that recipient African ancestry expressed as a quantitative variable (defined as R-pAFR) was associated with DCAL in a prospective renal transplant cohort.(22) While current concepts implicate altered tacrolimus metabolism,(18) specific induction and/or maintenance therapy,(19) and socioeconomic factors to account for these observations, these associations do not fully explain the worse outcomes in recipients with African ancestry.

As APOL1 G1/G2 alleles are seen exclusively in AAs and Hispanics (with recent African ancestry), herein we employed two prospective transplant cohorts(22, 23) to test for associations among the number of recipient APOL1 risk alleles (R-nAPOL1), R-pAFR and transplant outcomes. We report the unexpected association of R-nAPOL1 with DCAL in additive models, implying a role for even one APOL1 risk allele (either G1 or G2) in recipients, distinct from the previously reported association of two risk alleles in the donor with increased DCAL. This association was identified in all recipients as well as in AA and Hispanic (AA/H) recipients. We then identified a novel association of R-nAPOL1 with T cell-mediated rejection (TCMR), independent of recipient AA ethnicity, and validated these results externally. Finally, additional analyses implicate an unanticipated role for APOL1 risk alleles on immune activation, specifically in activated CD4^+^/CD8^+^ T cells and CD56^dim^ nature killer (NK) cells, providing a potential mechanism to account for the observed associations.

## RESULTS

### Study cohorts

The Genomics of Chronic Allograft Rejection (GOCAR)(24, 25) and Clinical Trials in Organ Transplantation-01/17 (CTOT-01/17, hereafter referred to as CTOT)(23) were prospective, multicenter, observational studies that enrolled crossmatch negative kidney transplant candidates. We employed a sub-cohort of 385 donor-recipient (D-R) pairs with genome-wide genotype data from GOCAR for discovery,(22) and a sub-cohort of 122 D-R pairs with genome-wide genotype data from CTOT as a validation set (see Materials and Methods; Figures S1 and S2). Demographic and clinicopathologic characteristics of GOCAR and CTOT cohorts, stratified by R-nAPOL1, are in Table 1 and published elsewhere.(22, 23) Clinical characteristics between the two cohorts are comparable (Table S1), although CTOT cohort contained a higher proportion of AA/H recipients and deceased donors. The GOCAR cohort had longer follow-up using UNOS and ANZDATA databases (mean follow-up 4.6 years) and thus more DCAL and TCMR events,(22) while CTOT collected information up to 5-years (mean follow-up 3.7 years). For each D-R pair from both cohorts, we used genome-wide genotype data excluding the MHC region(22) to estimate pAFR and infer genetic ancestry (Table S2). As expected, APOL1 genotyping showed that G1/G2 risk alleles are only detected in genetic AAs or Hispanics (i.e., AA/H) among D-Rs in both cohorts (see Methods; Tables S3 and S4). We observed a higher frequency of depletional induction agents in R-nAPOL1 subjects from both cohorts without differences in the number of D-R HLA mismatches between recipients carrying APOL1 risk alleles and non-carriers (Table 1).

**Table 1.** Demographic and clinicopathologic characteristics of donors and recipients in the GOCAR and CTOT cohorts stratified by the number of recipient APOL1 risk alleles.

| Variable | GOCAR<sup>a</sup> |  |  |  | CTOT<sup>b</sup> |  |  |  |
| --- | --- | --- | --- | --- | --- | --- | --- | --- |
|  | Recipient #<br>APOL1 risk<br>alleles = 0<br>(n = 316) | Recipient #<br>APOL1 risk<br>alleles = 1<br>(n = 20) | Recipient #<br>APOL1 risk<br>alleles = 2<br>(n = 20) | p-value<sup>c</sup> | Recipient #<br>APOL1 risk<br>alleles = 0<br>(n = 94) | Recipient #<br>APOL1 risk<br>alleles = 1<br>(n = 17) | Recipient #<br>APOL1 risk<br>alleles = 2<br>(n = 9) | p-value<sup>c</sup> |
| **Recipient characteristics** |  |  |  |  |  |  |  |  |
| **Death censored graft loss (years)** |  |  |  |  |  |  |  |  |
| mean ± SD; median (range) | 4.7 ± 1.6;<br>4.9 (0.04, 7.3) | 4.1 ± 2.0;<br>4.8 (0.3, 6.3) | 3.8 ± 1.9;<br>3.7 (0.8, 6.9) | **0.019** | 3.8 ± 1.7;<br>5.0 (0.02, 5.0) | 3.5 ± 1.9;<br>5.0 (0.02, 5.0) | 3.1 ± 2.1;<br>3.6 (0.1, 5.0) | 0.48 |
| # events (%) | 30 (9.4%) | 5 (25.0%) | 9 (45.0%) | **<0.001** | 2 (2.1%) | 2 (11.8%) | 2 (22.2%) | **0.02** |
| **TCMR ≥ borderline, # events (%)** | 103 (32.6%) | 7 (35.0%) | 8 (40.0%) | 0.75 | 8 (8.5%) | 3 (17.6%) | 3 (33.3%) | **0.05** |
| **TCMR > borderline, # events (%)** | 28 (8.9%) | 3 (15.0%) | 4 (20.0%) | 0.14 | 0 (0.0%) | 1 (5.9%) | 0 (0.0%) | 0.22 |
| **Recurrent TCMR ≥ borderline, # events (%)** | 46 (14.6%) | 4 (20.0%) | 6 (30.0%) | 0.15 | - | - | - | - |
| **Recurrent TCMR > borderline, # events (%)** | 17 (5.4%) | 3 (15.0%) | 4 (20.0%) | **0.02** | - | - | - | - |
| **Age (years), mean ± SD; median (range)** | 49.7 ± 13.9;<br>51 (18, 83) | 53.2 ± 10.4;<br>55 (24, 66) | 50.7 ± 12.6;<br>48 (27, 77) | 0.53 | 49.6 ± 13.5;<br>52 (18, 89) | 49.9 ± 12.5;<br>46 (35, 73) | 42.6 ± 14.6;<br>36 (22, 63) | 0.32 |
| **Gender, male, n (%)** | 213 (67.4%) | 14 (70.0%) | 11 (55.0%) | 0.55 | 58 (61.7%) | 10 (58.8%) | 5 (55.6%) | 0.95 |
| **Genetic ancestry<sup>e</sup>, n (%)** |  |  |  | **<0.001** |  |  |  | **<0.001** |
| African American | 12 (3.8%) | 16 (80.0%) | 15 (75.0%) |  | 8 (8.5%) | 14 (82.4%) | 8 (88.9%) |  |
| Hispanic | 56 (17.7%) | 4 (20.0%) | 5 (25.0%) |  | 12 (12.8%) | 3 (17.6%) | 1 (11.1%) |  |
| Asian | 13 (4.1%) | 0 (0.0%) | 0 (0.0%) |  | 2 (2.1%) | 0 (0%) | 0 (0%) |  |
| Caucasian | 235 (74.4%) | 0 (0.0%) | 0 (0.0%) |  | 72 (76.6%) | 0 (0%) | 0 (0%) |  |
| **HLA mismatch score<sup>d</sup>, mean ± SD** | 2.0 ± 1.0 | 2.3 ± 0.9 | 2.2 ± 0.8 | 0.30 | 3.1 ± 1.8 | 3.8 ± 1.3 | 4.1 ± 2.5 | 0.166 |
| **Induction, n (%)** |  |  |  | **<0.001** |  |  |  | **0.004** |
| No induction | 74 (23.4%) | 2 (10.0%) | 2 (10.0%) |  | 34 (36.2%) | 1 (5.9%) | 0 (0%) |  |
| Non-depletional (IL2 antagonist) | 122 (38.6%) | 3 (15.0%) | 2 (10.0%) |  | 26 (27.7%) | 4 (23.5%) | 5 (55.6%) |  |
| Depletional (Thymoglobulin or Campath) | 120 (38.0%) | 15 (75.0%) | 16 (80.0%) |  | 34 (36.2%) | 12 (70.6%) | 4 (44.4%) |  |
| **Donor characteristics** |  |  |  |  |  |  |  |  |
| **Age** (years), mean ± SD; median (range) | 42.5 ± 14.9; 45 (3, 73) | 44.8 ± 11.5; 48 (23, 60) | 42.6 ± 16.6; 45.5 (16, 73) | 0.80 | 40.6 ± 12.8; 41 (6, 62) | 37.3 ± 9.3; 37 (24, 59) | 40.8 ± 13.3; 39 (19, 65) | 0.59 |
| **Gender**, male, n (%) | 156 (49.4%) | 6 (30.0%) | 12 (60.0%) | 0.14 | 37 (39.4%) | 7 (41.2%) | 5 (55.6%) | 0.70 |
|  |  |  |  | **0.001** |  |  |  |  |
| **Genetic ancestry<sup>e</sup>, n (%)** |  |  |  |  |  |  |  |  |
|  |  |  |  |  |  |  |  | **<0.001** |
| African American | 11 (3.5%) | 2 (10.0%) | 6 (30.0%) |  | 10 (10.6%) | 11 (64.7%) | 5 (55.6%) |  |
| Hispanic | 39 (12.3%) | 4 (20.0%) | 4 (20.0%) |  | 11 (11.7%) | 3 (17.6%) | 2 (22.2%) |  |
| Asian | 6 (1.9%) | 0 (0.0%) | 0 (0.0%) |  | 2 (2.1%) | 0 (0%) | 0 (0%) |  |
| Caucasian | 260 (82.3%) | 14 (70.0%) | 10 (50.0%) |  | 71 (75.5%) | 3 (17.6%) | 2 (22.2%) |  |
| **Donor type**, live donor, n (%) | 172 (54.4%) | 7 (35.0%) | 5 (25.0%) | **0.01** | 83 (89.2%) | 14 (82.4%) | 6 (66.7%) | 0.12 |
| **Sub-cohort of AA/H recipients with additional molecular data<sup>f</sup>** |  |  |  |  |  |  |  |  |
| **Sample size**, n (%) | 34 (10.7%) | 14 (70%) | 12 (60%) |  | 15 (15.9%) | 3 (17.6%) | 2 (22.2%) |  |

<sup>a</sup>: Genome-wide genotype data is available for 385 donor-recipient (D-R) pairs from the parent GOCAR study.(22) There are missing data in the APOL1 genotype for 29 recipients and 14 donors (see Table S3 for details).
<sup>b</sup>: Genome-wide genotype data is available for 122 donor-recipient (D-R) pairs from the parent CTOT study. There are missing data in the APOL1 genotype for 2 recipients and 2 donors (see Figures S1 and S2 and Table S3 for details).
<sup>c</sup>: P-value was calculated from ANOVA for continuous variables and from Fisher's exact test for categorical variables unless otherwise specified.
<sup>d</sup>: HLA mismatch score was derived from 2-digit HLA allele typing. Following previous reports for GOCAR,(22, 24, 25) the raw mismatch score (scaling from 0 to 6) was categorized into: 0 (no mismatches), 1 (1-2 mismatches), 2 (3-4 mismatches), and 3 (5-6 mismatches); while for the CTOT cohort, the raw mismatch score (scaling from 0 to 6) was used. In subsequent statistical analyses, this variable was used as a numeric covariate in regression models. The p-value for this variable in the current table was derived from Kruskal-Wallis test.
<sup>e</sup>: Genetic ancestry was inferred from genome-wide genotype data and considered more accurate than self-reported race.(22)
<sup>f</sup>: See materials and methods for detailed description of the data.

### Recipient APOL1 G1/G2 alleles associate with graft loss

We investigated the association of R-nAPOL1 with DCAL for all recipients and in the AA/H strata in the GOCAR (discovery) and CTOT (validation) cohorts. Kaplan-Meier survival curve analysis (Figure 1) stratified by the number of APOL1 risk alleles (0, 1, or 2) showed clear differentiation among groups, with the number of risk alleles correlating directly with higher risk of DCAL in both cohorts (GOCAR cohort: log-rank p-value < 0.0001; CTOT cohort: log-rank p-value = 0.0075). The finding supports an additive effect of the APOL1 risk alleles in recipients on graft survival, i.e. each copy of the risk alleles increases the risk of graft loss, distinct from prior data.(3) We next adjusted for covariates previously shown to be associated with DCAL,(22) including genetic ancestry, induction therapy, and donor type, using a multivariable Cox regression analysis (Table 2). This analysis revealed that in GOCAR, R-nAPOL1 remained associated with DCAL in an additive manner (HR = 2.14 per copy of risk alleles; p-value = 0.006), independent of recipients’ genetic ancestry. Analysis of the CTOT cohort validated the results (Table S5). When we performed a meta-analysis including both cohorts (Figure 2), we observed the hazard ratio of each additional risk allele was 2.27 (95% CI: 1.41 ∼ 3.63; p-value = 0.0007). When we performed *sensitivity analysis* within the strata of AA/H recipients, we observed a similar pattern of significantly separated Kaplan-Meier survival curves for the three R-nAPOL1 groups (Figure S3A) as well as a significant association between R-nAPOL1 and DCAL with similar effect size (Table 2) in GOCAR. Within the AA/H strata of CTOT, we found that the pattern of differentiated survival curves (Figure S3B) and the positive association of R-nAPOL1 with DCAL (Table S5) remained, albeit with diminished significance level due to limited sample size. To account for the effect on allograft survival of donor APOL1 risk genotype, where high-risk genotype in donors is defined as 2 copies of G1/G2 alleles and low-risk genotype as 0 or 1 copy of G1/G2 allele, we performed additional sensitivity analyses stratified by donor APOL1 risk genotype in both the cohorts. The multivariable Cox regression analyses were performed on the stratum of donors carrying the low-risk genotype, as the sample sizes for the high-risk group in both cohorts are limited. The results from stratified analyses remain similar as in the main analysis: the R-nAPOL1 was associated with DCAL, independent of donor APOL1 risk genotype, for all recipients and for AA/H recipients (Tables S6 and S7). Together, these data demonstrate that R-nAPOL1 associates with DCAL in an additive manner in both cohorts.

**Figure 1.**
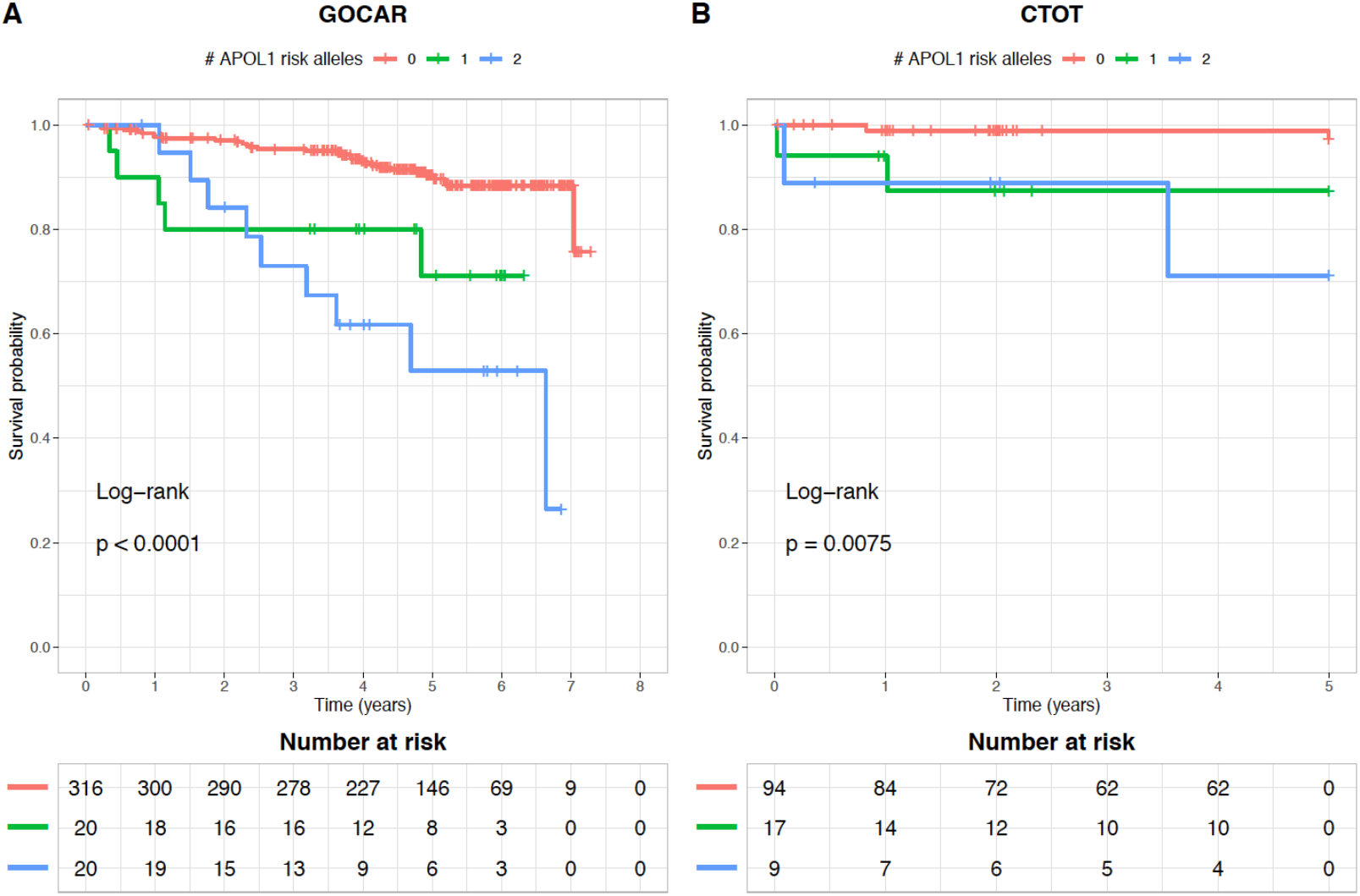
Kaplan-Meier plot of death-censored allograft survival for recipients with different numbers of APOL1 risk alleles. (**A**) GOCAR; (**B**) CTOT.

**Figure 2.**
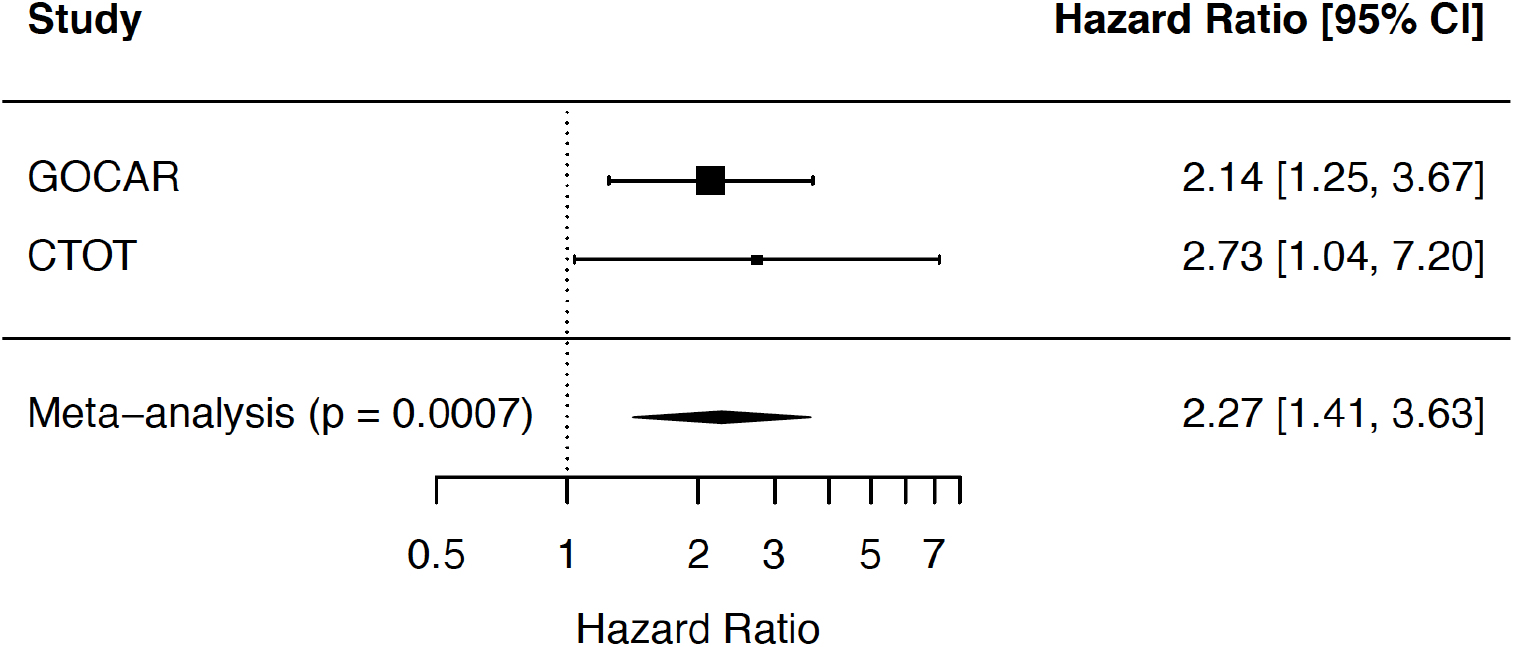
Meta-analysis for the association of APOL1 risk alleles with death-censored allograft survival across GOCAR and CTOT. Hazard ratio estimates of the number of APOL1 risk alleles on death-censored allograft loss (DCAL) from the GOCAR and CTOT cohorts for all ancestries were included in fixed effect meta-analysis. The hazard ratios and corresponding 95% confidence intervals (CI) for individual studies and meta-analysis were visualized by forest plot. The size of the squares shown for individual studies is proportional to the sample size of each study.

**Table 2.** Association of APOL1 risk alleles with death-censored allograft loss in an additive manner using multivariable Cox regression.

| **Variable** | **HR** | **95% CI** | **p-value** |
| --- | --- | --- | --- |
| ***GOCAR: recipients of all ancestries<sup>a</sup> (n = 343<sup>b</sup>; 44 [12.8%] graft loss events)*** |  |  |  |
| **# APOL1 risk alleles** | 2.14 | (1.25, 3.67) | **0.006** |
| **Recipient genetic ancestry (ref: Caucasian)** |  |  |  |
| **African American** | 0.96 | (0.29, 3.13) | 0.95 |
| **Hispanic** | 2.62 | (1.21, 5.70) | **0.01** |
| **Induction (ref: No)** |  |  |  |
| **Non-depletional** | 2.94 | (0.95, 9.13) | 0.06 |
| **Depletional** | 3.57 | (1.17, 10.9) | **0.03** |
| **Donor type (ref: LD) DD** | 2.57 | (1.27, 5.20) | **0.009** |
| **HLA mismatch score** | 1.26 | (0.87, 1.82) | 0.23 |
| ***GOCAR: recipients of African American and Hispanic (n = 108<sup>b</sup>; 26 [24.1%] graft loss events)*** |  |  |  |
| **# APOL1 risk alleles** | 2.32 | (1.33, 4.06) | **0.003** |
| **Recipient genetic ancestry (ref: AA)** | 3.06 | (1.03, 9.12) | **0.04** |
| Hispanic |  |  |  |
| **Induction (ref: No)** |  |  |  |
| **Non-depletional** | 6.22 | (0.72, 54.1) | 0.10 |
| **Depletional** | 6.03 | (0.75, 48.4) | 0.09 |
| **Donor type (ref: LD) DD** | 2.48 | (0.84, 7.26) | 0.10 |
| **HLA mismatch score** | 1.81 | (0.99, 3.33) | 0.06 |

<sup>a</sup>: The “Asian” category was excluded due to limited sample size which led to instable model fitting.
<sup>b</sup>: Sample size was reduced due to missing data in APOL1 risk alleles.

### Recipient APOL1 G1/G2 alleles associate with clinical and subclinical rejection

We next tested the strength of the association between R-nAPOL1 and TCMR episodes in the GOCAR and CTOT cohorts. The study designs captured clinical rejection episodes up to 2 years in both cohorts, as well as subclinical rejections at 3, 12, and 24 months (GOCAR), and at 6 months (CTOT).(23-25) In GOCAR, 126 recipients (32.7%) had at least one episode of subclinical or clinical TCMR (with Banff borderline or greater) identified among 3 surveillance biopsies,(22, 25) while in CTOT, 15 recipients (12.3%) had at least one TCMR episode including the 6-month surveillance biopsy.(23) R-nAPOL1 significantly associated with various TCMR outcomes in multivariable logistic regression models, independent of donor APOL1 risk genotype, with progressively increasing odds ratios present with more severe TCMR phenotypes (Table 3). For s*ensitivity analyses* in the subset of AA/H recipients, analyses showed that the association of R-nAPOL1 with different TCMR outcomes remained, with similar increasing odds ratio with increased severity of TCMR phenotypes. In the CTOT cohort, by logistic regression, we observed that the association of R-nAPOL1 with TCMR was significant for the whole cohort and the AA/H subset in univariate analysis, while the direction and magnitude of the association remained with reduced significance in multivariable analysis (Table S8). Taken together, these data indicate a strong association between R-nAPOL1 and TCMR events.

**Table 3.** Association of recipient APOL1 risk alleles with different TCMR outcomes using multivariable logistic regression in the GOCAR cohort.

| TCMR outcome<sup>a,b</sup> | n<sub>case</sub><sup>c</sup> | OR | 95% CI | p-value<sup>e</sup> |
| --- | --- | --- | --- | --- |
| **GOCAR: recipient of all ancestries (n<sub>control</sub> = 232)<sup>d</sup>** |  |  |  |  |
| **TCMR ≥ borderline** | 115 |  |  |  |
| Recipient # APOL1 risk alleles |  | 1.95 | (0.99, 3.95) | 0.06 |
| Donor APOL1 high-risk genotype |  | 0.54 | (0.02, 5.20) | 0.62 |
| **TCMR > borderline** | 34 |  |  |  |
| Recipient # APOL1 risk alleles |  | 2.74 | (1.10, 7.15) | **0.03** |
| Donor APOL1 high-risk genotype |  | 1.80 | (0.07, 24.2) | 0.67 |
| **Recurrent TCMR ≥ borderline** | 55 |  |  |  |
| Recipient # APOL1 risk alleles |  | 3.58 | (1.57, 8.78) | **0.003** |
| Donor APOL1 high-risk genotype |  | 0.75 | (0.03, 8.40) | 0.82 |
| **Recurrent TCMR > borderline** | 23 |  |  |  |
| Recipient # APOL1 risk alleles |  | 3.75 | (1.42, 10.7) | **0.009** |
| Donor APOL1 high-risk genotype |  | 1.59 | (0.05, 24.6) | 0.75 |
| **GOCAR: recipients of African American and Hispanic (n<sub>control</sub> = 66)<sup>d</sup>** |  |  |  |  |
| **TCMR ≥ borderline** | 35 |  |  |  |
| Recipient # APOL1 risk alleles |  | 1.98 | (0.99, 4.13) | 0.06 |
| Donor APOL1 high-risk genotype |  | 0.64 | (0.03, 7.31) | 0.74 |
| **TCMR > borderline** | 14 |  |  |  |
| Recipient # APOL1 risk alleles |  | 6.41 | (1.80, 34.3) | **0.01** |
| Donor APOL1 high-risk genotype |  | 1.22 | (0.01, 115.0) | 0.93 |
| **Recurrent TCMR ≥ borderline** | 19 |  |  |  |
| Recipient # APOL1 risk alleles |  | 3.45 | (1.44, 9.21) | **0.008** |
| Donor APOL1 high-risk genotype |  | 1.06 | (0.03, 18.2) | 0.97 |
| **Recurrent TCMR > borderline** | 12 |  |  |  |
| Recipient # APOL1 risk alleles |  | 7.78 | (2.09, 43.9) | **0.006** |
| Donor APOL1 high-risk genotype |  | 1.24 | (1.08, 98.1) | 0.92 |

<sup>a</sup>. In each multivariable logistic regression model, adjusted covariates include recipient genetic ancestry, center, induction, donor type, HLA mismatch score, and donor APOL1 high-risk genotype, where donor APOL1 high-risk genotype is defined as 2 copies of G1/G2 alleles and low-risk genotype as 0 or 1 G1/G2 allele. For concise presentation, only the results for the number of recipient APOL1 risk alleles and for donor APOL1 risk genotype were shown in the table.
<sup>b</sup>: TCMR outcomes include: (a) any Banff borderline or greater TCMR, (b) Banff 1A or greater TCMR, (c) recurrent (>1 episode) TCMR including borderline, and (d) recurrent TCMR with 1A or greater Banff score.
<sup>c</sup>: Sample size was reduced due to missing data in APOL1 risk alleles.
<sup>d</sup>: Controls (no TCMR) were defined as patients with either (a) no TCMR or borderline TCMR on obtained biopsies at anytime, or (b) no reported biopsies during follow up.
<sup>e</sup>: Bold p-value < 0.05.

### Recipient AA ancestry associate with creatinine levels up to 24-months post-transplant and not TCMR

Since we found a correlation between R-pAFR and R-nAPOL1, we tested for associations between R-pAFR and transplant outcomes independent of R-nAPOL1, noting that we previously reported that R-pAFR did not associate with Banff inflammation subscores or TCMR up to 2 years post-transplant.(22) This previous work also showed that no other Banff component scores in biopsies obtained within 2 years associated with R-pAFR in GOCAR. Herein, using GOCAR and CTOT cohorts, we used linear mixed models incorporating all available longitudinal creatinine data (to account for intra-individual variability) to determine the association between R-pAFR and creatinine levels (or estimated GFR) as a measure of kidney function. This analysis revealed that in GOCAR, R-pAFR significantly associated with serum creatinine levels from 3 to 24 months after transplant, independent of R-nAPOL1, post-transplant recipient BMI (to account for creatinine generation), donor APOL1 risk genotype, and donor pAFR (to account for AA-to-AA transplants) (Table 4; Figure S4). For example, as shown in Table 4, a recipient with 100% of African ancestry has on average 0.75 mg/dL higher serum creatinine than a recipient with 0% of African ancestry, or equivalently, every 10% increment of African ancestry in recipient would lead to a 0.075 mg/dL increment in creatinine level. We confirmed this association in CTOT with creatinine levels between 3 and 12 months after transplant (Table 4; Figure S4). Estimated GFR by modified MDRD or CKD-EPI equations(26) tended to inversely correlate with R-pAFR in mixed models but insignificantly (p-value = 0.06; not shown). These data relayed distinct post-transplant phenotypic associations of recipient African ancestry and recipient APOL1 risk allele status, in our study cohorts.

**Table 4.** Association of recipient pAFR with creatinine using linear mixed model.

| Variable<sup>a</sup> | Coefficient<br>(mg/dL) | 95% CI | P-value<sup>c</sup> |
| --- | --- | --- | --- |
| **GOCAR (n = 320 D-R pairs)<sup>b</sup>** |  |  |  |
| Recipient pAFR | 0.75 | (0.32, 1.19) | **<0.001** |
| Donor pAFR | -0.18 | (-0.76, 0.39) | 0.53 |
| Recipient # APOL1 risk alleles | -0.08 | (-0.31, 0.15) | 0.49 |
| Donor APOL1 high risk genotype | 0.19 | (-0.62, 1.00) | 0.65 |
| HLA mismatch score | -0.002 | (-0.08, 0.08) | 0.97 |
| Time (months) | 0.001 | (-0.001, 0.004) | 0.33 |
| **CTOT (n = 107 D-R pairs)<sup>b</sup>** |  |  |  |
| Recipient pAFR | 0.39 | (0.03, 0.75) | **0.03** |
| Donor pAFR | -0.12 | (-0.25, 0.01) | 0.07 |
| Recipient # APOL1 risk alleles | 0.03 | (-0.11, 0.17) | 0.66 |
| Donor APOL1 high risk genotype | -0.15 | (-0.48, 0.19) | 0.38 |
| HLA mismatch score | 0.02 | (-0.02, 0.05) | 0.34 |
| Time (months) | -0.002 | (-0.009, 0.004) | 0.50 |

<sup>a</sup>: In the multivariable linear mixed regression model, fixed-effect covariates include recipient age, sex, pAFR, number of APOL1 risk alleles, and BMI; donor age, sex, pAFR, and APOL1 high-risk genotype, where high-risk genotype is defined as 2 copies of G1/G2 alleles and low-risk genotype as 0 or 1 G1/G2 allele; induction, donor type, and HLA mismatch score. Subject-wise random effect was accounted for in the model. For concise presentation, only genetic relevant variables and time were shown in the table.
<sup>b</sup>: Sample size was reduced due to missing data in covariates.
<sup>c</sup>: Bold p-value < 0.05.

### SNP-based mismatches in APOL1 between donor-recipient pairs do not associate with DCAL

We further asked whether the association of R-nAPOL1 with DCAL was related to, or independent of, “mismatches” at the APOL1 locus itself, between D-R pairs. This was especially relevant since 90.3% (GOCAR) and 82.5% (CTOT) of donors had G0/G0 genotype, while 11.2% (GOCAR) and 21.7% (CTOT) of recipients had either one or two copies of G1 and/or G2 alleles (Table S3), increasing the likelihood of an APOL1 D-R mismatch among recipients with APOL1 risk alleles. We defined a SNP-based mismatch score to quantify the overall mismatch between any given D-R pair at the APOL1 locus, and to reflect the overall effect of the introduction of any new APOL1 variants from the donor kidney into the recipient (see Methods). Multivariable Cox models showed that APOL1 SNP-based mismatch score had no significant effect on DCAL (Table S9), and conditional on the APOL1 SNP-based mismatch score, the R-nAPOL1 remained associated with DCAL for all the recipients and the AA/H recipients in both cohorts (Table S9). This suggests an intrinsic effect of APOL1 risk alleles in recipients on DCAL independent of APOL1 D-R “mismatches”.

### Phenotypic data from immune cell types carrying APOL1 risk alleles in AA/H recipients show immune activation

Since R-nAPOL1, rather than mismatches at APOL1 locus between donors and recipients, associated with DCAL and TCMR, we examined immune cell phenotype and function in AA/H recipients with APOL1 risk alleles using auxiliary data from public datasets, GOCAR, and CTOT (see Methods). First, we confirmed APOL1 protein expression in peripheral blood mononuclear cells (PBMCs) using a discarded leukapheresis sample (Figure 3A). Positive and negative controls, respectively, included APOL1 over-expressing human podocytes and a mouse macrophage cell line (see Methods).

**Figure 3.**
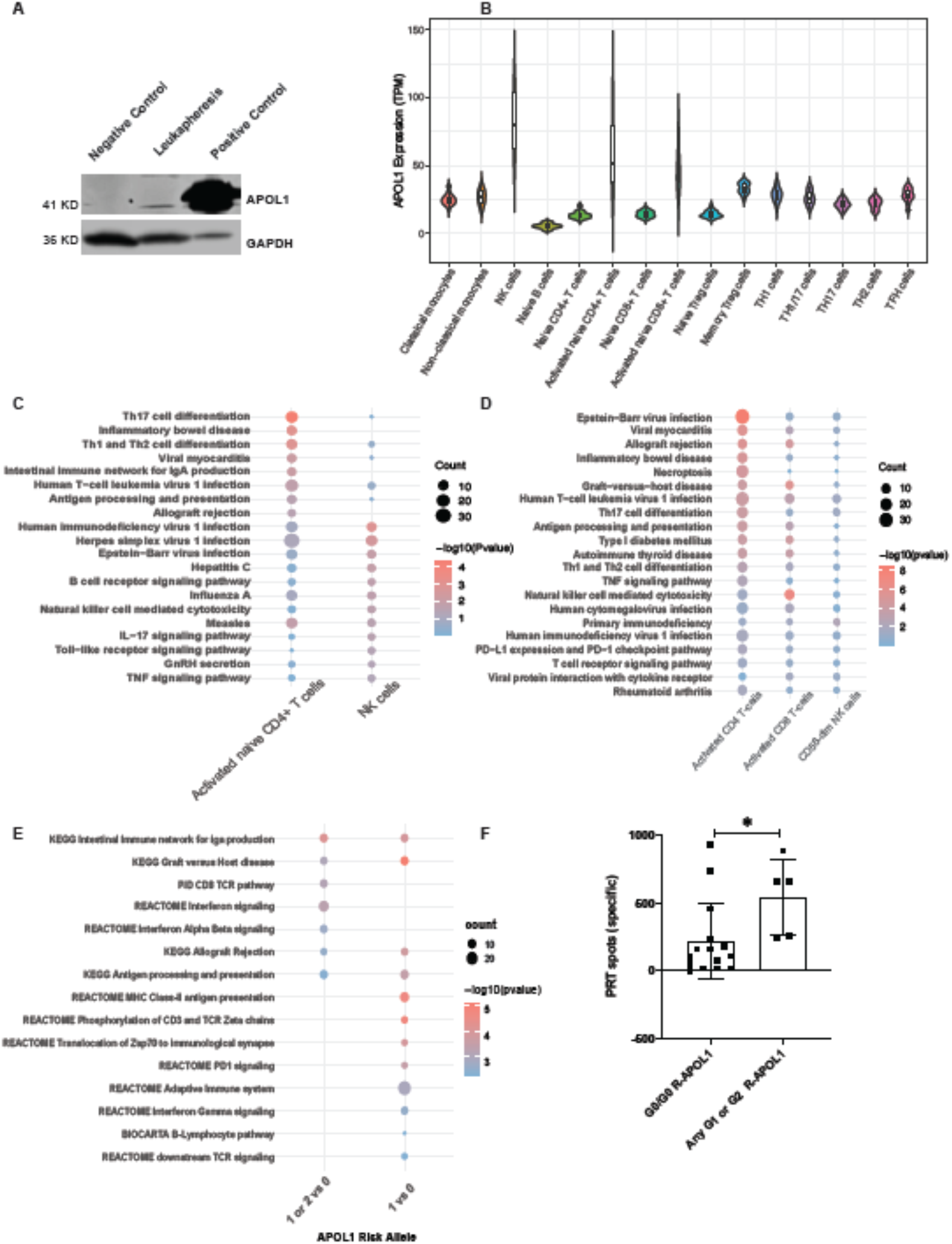
Phenotypic data from immune cell types carrying APOL1 risk alleles in AA/H recipients show immune activation. (**A**) APOL1 expression at protein level confirmed by western blot. (**B**) APOL1 mRNA expression of DICE AA/H individuals in 15 different types of immune cells. (**C**) Enrichment in immune related pathways of DEGs identified by comparing DICE AA/H individuals carrying one or two copies of G1 or G2 alleles vs. G0/G0 genotype in activated CD4^+^ T cells and CD56^dim^ NK cells. (**D**) Enrichment in immune related pathways of DEGs identified by comparing single cell RNA sequencing data of two AA/H ESRD patients with G2/G0 genotype vs. G0/G0 genotype in activated CD4^+^ and CD8^+^ T cells and CD56^dim^ NK cells. (**E**) Enrichment of DEGs in immune related pathways when comparing GOCAR AA/H recipients carrying one or two copies of G1 or G2 alleles vs. G0/G0 genotype, or comparing those carrying any one risk allele vs. G0/G0 genotype. (**F**) Panel reactive T cell ELISPOT assay comparing CTOT AA/H recipients with any APOL1 G1 or G2 allele vs. G0/G0 genotype. *: Wilcoxon rank-sum test p-value < 0.05. DEG: differentially expressed gene; AA/H: African American and Hispanic.

Since PBMCs include a mixture of mononuclear cells, to understand cell type specific expression of APOL1, we utilized the data generated by the DICE project,(27) where bulk RNA sequencing data for 15 sorted immune cell types and APOL1 genotype information was available for 91 healthy individuals. We focused on the subset of 22 AA/H individuals, five of whom carried one or two copies of G1/G2 alleles. Among the 15 cell types, we discerned that APOL1 mRNA expression was highest in CD56^dim^ NK cells, and *ex vivo* polyclonally activated CD4^+^ and CD8^+^ T cells (but not in unstimulated T cells, Figure 3B). We next performed differential gene expression analysis to identify differentially expressed genes (DEGs) in individuals with any G1/G2 alleles as compared to those with G0/G0 genotype (see Methods). Our analyses showed significant enrichment of DEGs in pathways involved in immune activation within activated CD4^+^ T cells and cytotoxic CD56^dim^ NK cells from subjects with any vs. no G1/G2 alleles (Figure 3C; Table S10). Within activated CD4^+^ T cells from these healthy controls with APOL1 risk alleles, we observed enrichment of genes involved in allograft rejection and antigen processing pathways (HLA genes), T cell activation and differentiation (*IL2, IL21, IL21R, IL18R, GATA3*), and chemokine genes (*CXCL8, CXCL3, CXCL11*). In both CD4^+^ lymphocytes and cytotoxic NK cells, DEGs included TNF**α**-signaling pathway genes and antiviral response genes (Table S10).

To further investigate transcriptomes of NK cells and activated T cells, we utilized our groups’ single cell RNA sequencing (scRNAseq) data generated from PBMCs of two ESRD patients listed for transplantation (GEO accession: GSE162470; see Methods). We used raw scRNAseq reads to call variants at APOL1 locus (see Methods), identifying one patient as G2/G0 genotype, while the other had G0/G0 genotype (Figure S5A), and simultaneously confirmed the expression of APOL1 G2 risk allele mRNA in PBMCs (Figure S5B). In the scRNAseq data, we confirmed enrichment of DEGs in immune related pathways in activated CD4^+^ and CD8^+^ T cells as well as in CD56^dim^ NK cells (Figure 3D; Table S11). In the single cell transcriptome of activated CD4^+^ T cells from the G2/G0 waitlisted patient, similar to healthy controls, we identified significant enrichment of DEGs associated with allograft rejection, antigen processing, and graft-vs-host disease. In APOL1 risk allele-carrying NK cells and stimulated CD4^+^ T cells from DICE data, as well as in activated CD8^+^ T cells in scRNAseq data, we observed upregulation of *IFNG* transcripts (Table S11).

To validate the immune activation signature of APOL1 risk genotypes in our cohorts, we examined pre-transplant peripheral blood transcriptomes of AA/H recipients within GOCAR.(28) In GOCAR, APOL1 genotyping and pre-transplant blood transcriptomes were available for 60 recipients (Table 1), who had one or two copies of G1/G2 alleles (n = 26), or G0/G0 genotype (n = 34). We performed differential gene expression analysis on whole blood mRNA samples prior to transplantation (see Methods).(28) Interestingly, recipients with any copy of G1 or G2 alleles showed DEGs enriched in immune response pathways as compared to G0/G0 recipients (Figure 3E; Table S12). We identified DEGs associated with allo- and antiviral-response pathways, similar to the DICE and scRNAseq data, as significantly enriched in peripheral transcriptomes of GOCAR recipients with 1 or 2 copies of the risk alleles (Table S12).

In an effort to assess whether APOL1 risk alleles associated with T cell function we re-analyzed ELISPOT data from the CTOT cohort in which AA/H recipients with APOL1 genotype information were tested for frequencies of alloreactive IFNg-producing PBMC co-cultured with a panel of six, HLA-disparate, stimulator cell lines.(29) These analyses showed stronger responses in AA/H recipients with any APOL1 risk alleles (n = 5) vs. G0/G0 recipients (n = 15) (Table 1; Figure 3F).

## DISCUSSION

Using two large prospective kidney transplant cohorts, we show for the first time that recipient APOL1 risk alleles are associated with long-term death-censored graft survival, and clinical/subclinical as well as recurrent TCMR events up to two years after transplantation. These findings were identified in additive models of APOL1 genotype, showing that even single risk allele in recipients presents increased risk for both acute rejection and long-term graft survival. We then used *in silico* and *ex vivo* auxiliary data from immune cells to confirm the expression of APOL1 at mRNA and protein levels in PBMCs, and demonstrate enrichment of pathways involving generic immune responses, as well as interferon-gamma ELISPOT responses. We identified cell types with higher APOL1 expression utilizing the DICE data (RNA sequencing) from healthy individuals(27) --CD56^dim^ NK cells, and *ex vivo* stimulated CD4^+^ and CD8^+^ T cells. In stimulated CD4^+^ T cells, significant enrichment of immune response pathways were confirmed among DICE participants with one or two copies of G1 or G2 alleles vs. G0/G0 genotype. Activated CD4^+^ and CD8^+^ T cells from scRNAseq data from a G2/G0 ESRD patient also showed consistent findings. Together these data support a novel role for FSGS-associated G1/G2 APOL1 alleles in immune cells in modulating allo-immune responses.

While the association of APOL1 G1/G2 risk alleles with the lifetime risk of ESRD and FSGS in AAs and admixed populations with African ancestry has been repeatedly affirmed in clinical data,(3, 4) data regarding the mechanism of adverse impact has been focused on the expression of mutant APOL1 protein,(5, 6) or mRNA in kidney epithelial cells.(30) In renal transplantation, the association of donor APOL1 risk alleles with DCAL have been consistently observed in retrospective data,(8-10) possibly via allograft FSGS.(14) Similar large-scale examinations of the association of recipient APOL1 risk allele with graft outcomes have not been reported. A single center retrospective study of 119 African American renal recipients did not find the association of recipient APOL1 risk allele with DCAL.(15) However, donor APOL1 genotypes were unknown here. Further, this study reported an unusually high DCAL rate of 25% at 5 years (vs. 5% and 11% for living- and deceased-donor kidneys in recent SRTR data), which likely contributed to inability to identify significant impact of R-nAPOL1 in this dataset.(17)

An issue with APOL1 association studies in transplantation is the exclusive association of G1/G2 alleles with African ancestry, a potential confounder for transplant outcomes.(30) In our data we addressed this issue in adjusted models using genetic ancestry (which was previously reported as more accurate than self-reported ancestry by our group (22) and others (31)) or R-pAFR (a quantitative measurement of ancestry), inferred from genome-wide genotype data, as well as in the strata of AA/H recipients, strengthening our findings of the association of APOL1 risk alleles with DCAL. We then identified a novel association of R-pAFR (and not R-nAPOL1) with serum creatinine levels within 2 years post transplantation. Furthermore, adjustment for longitudinal creatinine levels as a time-varying covariate in the survival models did not significantly attenuate the association of R-nAPOL1 with DCAL suggesting creatinine levels were not a mediator for the association of R-nAPOL1 with DCAL (not shown). Since transplant recipients are dependent on the allograft for creatinine excretion, the novel association between R-pAFR and post-transplant creatinine levels, may reflect increased generation of creatinine in recipients with African ancestry. Hence, our data reflective of genetically determined changes in creatinine levels are also timely to the ongoing discussion of the role of ancestry adjustment in estimated GFR equations,(32) and contributory to the use of genetic ancestry for these purposes.(33)

Our data raise new avenues for the investigation of APOL1 in allo-immune responses in renal transplantation --the role of APOL1 gene products in CD4^+^ and CD8^+^ T cells and cytolytic NK cells (CD56^dim^), the role of wild type vs. variant APOL1 protein/mRNA, and gain-of-function vs. loss-of-function mechanisms -- all need to be comprehensively examined. While renal epithelial cell injury mechanisms from APOL1 risk variants have been the subject to intense study,(5, 6, 30) APOL1 homologues were originally identified as TNFα responsive genes in endothelial cells.(34, 35) In humans, the APOL1 promoter has binding sequences for STAT2 and interferon responsive transcription factors, and a role for APOL1 as a cellular immune response gene in antiviral immunity has been postulated to explain its association with HIV-associated nephropathy.(36) Collapsing FSGS after viral infection was reported in APOL1 risk allele-carrying allografts,(14) and recently in a recipient with APOL1 risk allele after COVID-19 infection.(37) In lupus nephritis, where APOL1 risk genotypes associate with increased progression of disease, a toxic gain-of-function role for APOL1 variants by disrupting T cell autophagy and interferon signaling has been postulated.(38) Consistently, progressive nephritis was worsened with every copy of risk allele in Brazilian AA/H lupus patients.(39) In this context, our data implicating a role for APOL1 within T cells involved in adaptive immune responses against a donor organ, demonstrate that previous data regarding R-nAPOL1 and allograft outcomes need to be reinterpreted, and its role in infiltrating inflammatory mononuclear cells in native kidney glomerulonephritis investigated.

While our data provide new insights, we acknowledge several limitations. First, while we adjusted for biologic confounders based on clinical data collected in both cohorts, we cannot eliminate residual confounding effects of other factors including socio-economic and behavioral data (e.g., non-adherence), which were not collected. Second, in the CTOT cohort, our validation efforts for graft loss and acute rejection in AA/H sub-cohort did not reach statistical significance due to limited sample size and a paucity of events, although the effect sizes observed were similar. Third, the sample size of AA/H donors with African ancestry in both cohorts was limited, leading to few observed APOL1 risk alleles in donors. Larger multi-ethnic cohorts with adequate sample size of APOL1 risk alleles in D-R pairs will help evaluate the interaction of APOL1 risk alleles in donor organs and recipients on long-term transplant outcomes.(16) Last, the results from DEGs and enrichment analysis were only at nominal significance level given the limited sample sizes and the burden of multiple hypothesis testing. Nevertheless, rather than drawing conclusive results, we aimed to find suggestive directions in terms of pathways and cell types where APOL1 risk alleles may have effects on transcriptome and the transplant outcomes. Our findings are a platform to investigate cell-type specific immune functions of APOL1 in novel experimental models such as the human BAC transgenic mouse strains expressing either G0, G1 or G2 genes at physiological levels while remaining responsive to endogenous cytokine stimuli.

In summary, using two prospective transplant cohorts, we report for the first time the association of recipient APOL1 risk alleles with allograft survival and cellular rejection events. We demonstrate these associations in additive models showing the role of even single copy of G1/G2 alleles in the observed outcomes. We show phenotypic data supporting immune effects of APOL1 expression in specific cell types. We also report the association of African ancestry in recipients, quantified as R-pAFR, with serum creatinine post transplant. Our work forms a basis for further mechanistic work to understand the immunologic role of APOL1.

## MATERIALS AND METHODS

### Discovery cohort

The Genomics of Chronic Allograft Rejection (GOCAR) study is a prospective, multicenter study to examine the utility of genomics and genetics to predict the development of chronic allograft injury. Patients included in the study were prospectively enrolled from May 12, 2007, to July 30, 2011. Details of the study were reported elsewhere.(22, 24, 25) Enrolled patients had clinical data and lab draws at baseline, 3, 12, and 24 months after renal transplantation.

### Validation cohort

The Clinical Trials in Organ Transplantation-01/17 (CTOT-01/17, hereafter referred to as CTOT) was a prospective, multicenter, observational study that enrolled crossmatch negative kidney transplant candidates with 2 years follow up.(40) Adult and pediatric subjects undergoing a primary kidney transplant and having a negative flow cytometry crossmatch at the time of transplantation were eligible for enrollment. In the current study, only adult subjects with age >= 18 years and graft survival more than at least one week were included. Plans for multi-organ transplantation and/or clinically significant liver disease were exclusion criteria. The overall objective of CTOT-01 was to determine the relationships between results of immune assays and a composite primary endpoint (clinically evident or subclinical biopsy-proven cellular acute rejection with Banff grade 1A or higher, increase in Banff chronic sum score > 2, increase in interstitial fibrosis > 15%, graft loss, or death at 6 months after transplant) and/or a change in renal function (>30% decrease in estimated GFR [eGFR]) between 6 and 24 months after transplant. CTOT-17 (extension study of CTOT-01) was designed to collect information on 5-year outcomes in this cohort. Details of this cohort have been published previously.(23)

### Genotyping, data processing, and quality control

The genotyping and quality control (QC) for the GOCAR cohort has been reported previously.(22) After data processing and QC, complete genotype-phenotype data for 385 D-R pairs and 131,035 SNPs remained for statistical analysis. We applied the same procedure as done for GOCAR to CTOT. Briefly, recipient DNA was obtained from peripheral blood mononuclear cells (PBMCs), while donor DNA was obtained from either pre-perfusion allograft biopsies (in deceased donors, DDs) or PBMCs (in living donors, LDs). In the case where DNAs from both sources were available, the genotype data was derived from PBMC DNA. Illumina Infinium Global Screening Array (GSAMD-24v1-0_20011747_A1) was applied on the extracted DNA. The raw genotype data was subject to a series of QC steps (Figure S1). In sample-wise QC, we excluded samples based on criteria: 1) genetically inferred gender inconsistent with reported gender; 2) missing genotype rate > 0.03; 3) excessive genome-wide heterozygosity (indicating potential sample contamination); 4) individuals with European ancestry but carrying APOL1 G1/G2 risk alleles (see APOL1 genotyping section). In SNP-wise QC, we excluded SNPs based on criteria: 1) missing rate > 0.05; 2) minor allele frequency (MAF) < 0.01; 3) Hardy-Weinberg equilibrium (HWE) p-value < 1e-6. The markers with no chromosome information, or with ambiguous alleles (A/T or C/G), or not located on autosomes were excluded as well.

To prepare for downstream analysis (see ADMIXTURE analysis section), the processed genotype data from CTOT samples were merged with the genotype data from 1000 Genomes Project (KGP)(41) samples at shared SNP loci on autosomal chromosomes. From merged data, common SNPs with MAF > 0.05 were selected, where MAF was estimated based on KGP samples. The list of SNP markers in high density was pruned based on pairwise linkage disequilibrium,(42) where pairwise linkage disequilibrium between SNPs was derived from KGP samples. In order to explore the genetic effect beyond HLA, we excluded SNPs located in the MHC region in subsequent genetic analyses. After these steps, there were 122 D-R pairs with complete genotype data and 126,872 SNPs left in the CTOT cohort (Figure S1).

### ADMIXTURE analysis and genetic-inferred ancestry

We have used ADMIXTURE(43) to estimate the proportions of genetic ancestries of donors and recipients and infer their genetic ancestries for the GOCAR cohort as previously detailed.(22) The same analysis pipeline was also applied to the processed genotype data of the CTOT cohort. Briefly, we applied ADMIXTURE on the genome-wide genotype data with 1000 Genomes Project (KGP) Phase I(41) as reference populations to anchor the major ancestral populations. The genetic background of each individual was inferred as a mixture of four ancestral components, corresponding to African, Caucasian, East Asian and Native American ancestry (Figure S2). As shown in Figure S2A, the estimated proportions of African (pAFR) and Caucasian (pEUR) ancestry were used to define, in a conventional meaning, the genetic ancestry of donors and recipients. With simple thresholds, the individuals were categorized as African American if pAFR >= 0.6, Caucasian if pEUR >= 0.9, Asian if pAFR + pEUR <= 0.1 (and proportion of East Asian (pASN) >= 0.9), and Hispanic (i.e. admixed population with a spectral mixture of Caucasian, African, and Native American ancestral components) otherwise (Figure S2B).

### APOL1 genotyping

The G1 allele of APOL1 is represented by rs73885319 and rs60910145, two missense SNPs in almost perfect linkage disequilibrium, while the G2 allele is represented by a 6-bp microdeletion rs143830837 (or equivalently rs71785313).(2) The allele that does not carry any of these variants is named G0 hereafter. In the GOCAR cohort, the three allele-representing markers were imputed by the SHAPIT(44) and IMPUTE2(45) pipeline (see section below). To ensure the quality of imputation, the posterior probability of imputed genotype was required to be greater than 0.95; otherwise, the imputed genotype was considered as missing data. Among the 385 D-R pairs with genotype information, there were missing data in APOL1 genotype for 29 recipients and 14 donors (Table S3). In the CTOT cohort, fortunately, the two representative variants rs73885319 and rs71785313 were genotyped directly by the SNP array platform used, and thus the APOL1 genotype could be defined accordingly. The individuals genetically determined as Caucasian but carrying G1/G2 alleles, contradictory to the origin of the risk variants from African ancestry, were excluded (Figure S1). Actually, some of the ancestry of origin-inconsistent APOL1 genotypes were later confirmed as genotyping errors by PCR. Among the 122 D-R pairs with genotype information, there was missing data in APOL1 genotype for 2 recipients and 2 donors due to failure in genotyping effort at these two variants (Table S3).

### APOL1 SNP-based mismatch score

We evaluated the SNP-wise mismatches at APOL1 locus for both cohorts following the similar procedures as described in recent literature.(46) First, genome-wide genotype imputation was performed for both cohorts. For GOCAR, the imputation was done by the SHAPIT(44) and IMPUTE2(45) pipeline with the 1000 Genomes Project Phase I data(47) as reference panel; while for CTOT, the imputation was done by the Michigan Imputation Server(48) (https://imputationserver.sph.umich.edu) with the Haplotype Reference Consortium (HRC) reference panel (Release 1.1).(49) Second, at each SNP locus, a mismatch score of 1 for a donor-recipient (D-R) pair was defined when the donor introduced any allele(s) that did not appear in the recipient, and 0 otherwise. Third, the SNP-wise mismatch scores of SNPs within the range of APOL1 locus (Chr 22: 36649117-36663577) were summed as a measure of total mismatch at the APOL1 locus, and then the raw values of APOL1 mismatch score for each D-R pair was normalized by the interquartile range (IQR) across D-R pairs within each cohort.

### PBMC RNA sequencing data analysis for a subgroup of GOCAR patients

The details of PBMC isolation for a subgroup of GOCAR patients for RNA sequencing experiments and data analysis pipeline have been reported by our group before.(28) Briefly, total RNA was extracted from whole blood drawn from transplant recipients before transplant, and mRNA sequencing was performed on an Illumina HiSeq 4000 sequencer. Gene expression data was obtained from the GEO database (accession ID: GSE112927). In this study, we focused on the subgroup of 60 AA/H patients with genotype information available as well. Differential gene expression analysis was carried out by an R package limma(50) in comparison of recipients carrying one or two copies of APOL1 risk alleles (n = 26) vs. zero copy (n = 34), and in comparison of those with one copy (n = 14) vs. zero copy (n = 34). Differentially expressed genes (DEGs) were initially identified at p-value < 0.05. Biological functional pathways enriched for DEGs were determined by Fisher’s exact test at p-value < 0.05 using the information of biological process category in Gene Ontology (GO)(51) and pathways curated in the several pathway databases (KEGG,(52) Ingenuity IPA [QIAGEN Inc., https://www.qiagenbioinformatics.com/products/ingenuity-pathway-analysis], BIOCARTA,(53) Panther(54), PID(55), REACTOME(56), WikiPathways(57)).

### Panel reactive T cell assay for a subgroup of CTOT patients

Details and standardization of the IFNγ ELISPOT assay have been published by our group before.(40, 58) IFNγ production by recipient PBMCs against isolated *ex vivo* stimulated B cells from the respective donor, randomly chosen third party, and a standardized 6-donor panel were evaluated before transplant. Results were respectively reported as donor-specific, third-party, and PRT assay. ELISPOT data in a subgroup of CTOT AA/H recipients with one or two copies of APOL1 G1/G2 alleles (n = 5) were compared with those with G0/G0 genotype (n = 15). Experiment procedures are briefly described as follows. Blood samples from recipients were collected in heparinized green top tubes and PBMCs were isolated by Ficoll separation at each site within 6 hours of collection, and frozen using a standard operating procedure. Blood samples were obtained from living donors and processed similarly. PBMCs or spleen cells obtained from deceased donors were sent to the Mount Sinai core laboratory where they were processed and frozen. Recipient PBMCs (300,000 per well) were stimulated against respective stimulator cells (100,000 per well) in triplicate. The resulting spots were counted with an Immunospot computer-assisted ELISPOT image analyzer (Cellular Technology, Cleveland, OH). Results were depicted as the mean number of IFNγ spots per 300,000 recipient peripheral blood lymphocytes based on duplicate or triplicate measurements in a given assay.

### DICE RNA sequencing data analysis

To explore the expression of APOL1 risk alleles and associated gene signatures in various immune cell types, we utilized the RNA sequencing data generated by the DICE (Database of Immune Cell Expression, Expression quantitative trait loci [eQTLs] and Epigenomics) project (https://dice-database.org/). Access to the DICE data located on the dbGaP was requested (request #97206-2) and approved (dbGaP study accession number: phs001703). The description of the dataset has been detailed in literature.(27) Briefly, whole transcriptomic data was generated by bulk RNA sequencing from immune cell types isolated from leukapheresis samples provided by 91 healthy subjects. The cell types surveyed include three innate immune cell types (CD14^high^ CD16^-^ classical monocytes, CD14^-^ CD16^+^ non-classical monocytes, CD56^dim^ CD16^+^ NK cells), four adaptive immune cell types (naive B cells, naive CD4^+^ T cells, naive CD8^+^ T cells, and naive regulatory T cells [T_REG_]), six CD4^+^ memory or more differentiated T cell types (T_H_1, T_H_1/17, T_H_17, T_H_2, follicular helper T cell [T_FH_], and memory T_REG_), and two activated cell types (naive CD4^+^ and CD8^+^ T cells that were stimulated *ex vivo*).(27) In this study, the analysis was mainly focused on a subgroup of 22 AA/H individuals. The gene expression data was measured as transcript per million reads (TPM). Genes with mean TPM less than 1 across all samples were excluded from further analysis. Raw TPM expression profiles were log2-transferred by log2(TPM + 1), where a value of 1 was added to account for 0 values in TPM. Differentially expressed genes (DEG) between the group of individuals with one or two copies of APOL risk alleles (n = 5) and the group of individuals without any risk alleles (n = 17) were identified using the functions “contrasts.fit” and “eBayes” implemented in an R package limma (version 3.38.3).(59) Genes with p-value < 0.05 were considered to be nominally significant. Pathway enrichment analysis for DEGs was performed using clusterProfiler,(60) based on the KEGG pathway database.(52) P-value < 0.05 in enrichment analysis was considered nominally significant. Adjusted p-value using Benjamini-Hochberg (BH) method(61) and a q-value quantifying false discovery rate (FDR) using an R package qvalue(62) were also provided for multiple hypothesis testing control.

### Single cell RNA sequencing data analysis for two ESRD patients

We utilized single cell RNA sequencing (scRNAseq) data from PBMC samples collected from two ESRD patients (GEO accession: GSE162470). These data were downloaded and subjected to the pipeline as follows. Raw scRNAseq data was aligned using CellRanger (version 3.1.0) (10x Genomics, https://support.10xgenomics.com/single-cell-gene-expression/software/pipelines/latest/what-is-cell-ranger) and cells were filtered and clustered using Seurat (version 3.1.5) with default parameters.(63) Cell types were identified using classic immune markers as described in other PBMC studies (Table S13).(64) Short reads generated from the two single cell samples were aligned to human reference genome (GRCh37) by STAR(65) and genotyped following the GATK best practice for RNA sequencing data.(66) G1 and G2 alleles were identified based on the genotyped variants described in literature.(22) DEGs between two patients in each cell type were identified using the *FindMarkers* function from the Seurat package, and genes with Wilcoxon rank-sum test p-value < 0.01 were considered to be nominally significant. Pathway enrichment analysis for DEGs were performed the same way as described above for the DICE data analysis.

### APOL1 over-expression podocytes

Human Apol1 was amplified using cDNA synthesized with human podocyte mRNA as template by PCR. FLAG peptide sequence was incorporated into the antisense primer containing a terminal Xbal site. The sense primer contained a terminal EcoRV site.

Forward primer sequence:

<u>GATATCatggagggagctgctttgctgagag.</u>

Reverse primer sequence:

<u>tctagatcaCTTGTCGTCATCGTCTTTGTAGTCcagttcttggtccgcctgc</u>.

PCR-amplified products were cloned into pGEM-T-vector (Promega A3600). Apol1 sequence was confirmed by DNA sequencing using T7 primer. cDNAs of Apol1 were released from T-vectors with EcorV and XbaI restriction enzymes and inserted into EcorV and XbaI-digested PCDNA4B vectors. Lentiviral transduction and stably infected human podocyte lines were created. APOL1-Flag expression was confirmed. As negative controls for APOL1 expression, we used lysates from a mouse macrophage cell line since mouse cells do not express APOL1.

### Western Blotting

PBMC from leukapheresis sample, or podocyte cell lines were lysed with a buffer containing 25 mM Tris-HCl pH 7.4, 150 mM NaCl, 1 mM EDTA, 1% NP-40 and 5% glycerol, a protease inhibitor mixture and tyrosine and serine/threonine phosphorylation and phosphatase inhibitors. Lysates were subjected to immunoblot analysis --APOL1 antibody (ABCAM [ab231523]), Rabbit polyclonal Anti GAPDH antibody (#5174, Cell Signaling).

### Statistical analysis

Descriptive statistics (mean, standard deviation [SD], median, and range) were used to summarize the baseline characteristics of the GOCAR and CTOT cohorts. When comparing the baseline characteristics between groups of recipients carrying different numbers of APOL1 risk alleles, Fisher’s exact test was used to calculate p-value for categorical variables, analysis of variance (ANOVA) for continuous variables, and Kruskal-Wallis test for ordinal variables. Kaplan-Meier plot was used to visualize and compare the death-censored graft survival curves between groups of recipients carrying different numbers of APOL1 risk alleles and a log-rank test was used to calculate the p-value. The association of time-to-event outcome (DCAL) with risk factors was evaluated with Cox regression. The association of dichotomous outcomes (different TCMR outcomes) with risk factors was evaluated with logistic regression. The association of the longitudinal creatinine levels with risk factors was evaluated with linear mixed models, implemented in the R package lme4.(67) The fixed-effect meta-analysis of GOCAR and CTOT results were implemented in the R package metafor.(68) In each regression analysis, the samples with missing data in relevant covariates were omitted. Two-sided p-value < 0.05 was considered as statistical significance unless otherwise specified. These statistical procedures were implemented in R.(69)

### Study approval

GOCAR study: Informed written consent was obtained from all study participants from the individual clinical sites at the time of enrollment into the original study protocol. IRB approval was obtained from all participating institutions. CTOT01/17 study: Informed written consent was obtained from all study participants from the individual clinical sites at the time of enrollment into the original study protocol. IRB approval was obtained from all participating institutions. Consent included utilization of de-identified genetic data for research purposes and retrospective data reporting.

## Data Availability

Gene expression data from a subgroup of GOCAR patients was obtained from the GEO database (accession ID: GSE112927).
Single cell RNA sequencing data from PBMC samples collected from two ESRD patients was obtained from GEO database (accession ID: GSE162470).
DICE data was obtained from dbGaP (study accession number : phs001703).

## AUTHOR CONTRIBUTIONS

Research/study design: ZZ, JCH, POC, PSH, BM and MCM; Bioinformatics analyses: ZS, ZZ, ZY, KH, HC, and WZ. Experimentation: QL, RL, and CW. Sample preparation/processing: KB, MP, and JF. Reagents: MCM, PSH, CJH, SI, BM. Clinical data analyses: KC, SGC, ZZ, ZS, MCM. Histopathology: FS, IWG, and RBC. Manuscript: ZZ, ZS, PSH and MCM. All authors contributed to editing of the manuscript.

## ACKNOWLEDGEMENTS

We thank all the patients, donors and their families, the participating clinical sites, and investigators in the GOCAR and CTOT-01/17 studies. MCM and ZZ acknowledge the Translational Collaborative Research Initiative Grant, “Non-HLA donor-recipient differences and allograft survival,” Department of Medicine, and the computational resources and staff expertise provided by Scientific Computing at the Icahn School of Medicine at Mount Sinai. MCM acknowledges funding from American Heart Association (AHA 15SDG25870018), NIH DK122164, and pilot funding from the CTOT-19 study (PI: Peter Heeger; NIH U01AI063594) to study non-HLA donor-recipient genetic differences. The data reported here are substudies of the GOCAR study (PI: B. Murphy) supported by NIH U01AI070107-03 and the CTOT-01 study (PI: P. Heeger) supported by NIH U01AI63594-06. KH is partially supported by NIEHS 1R01ES029212-01, NIA AG058635, NIDDK U24DK062429 and DK106593.

Funding for the study of “Impact of genetic polymorphisms on human immune cell gene expression” (DICE data) was provided from the NIAID R24-AI108564. The datasets used for the analyses described in this manuscript were obtained from dbGaP through dbGaP study accession number phs001703. Data from the study were provided by Pandurangan Vijayanand on behalf of his collaborators at La Jolla Institute for Allergy and Immunology.

