## Supplemental Materials for "Recipient APOL1 risk alleles associate with death-censored renal allograft survival and rejection episodes"

**Supplementary Figures**


| | |
|---|---|
| Figure S1. Flowchart of genotyping and quality control for CTOT cohort | 1 |
| Figure S2. Genetic ancestry of CTOT donors and recipients | 2-3 |
| Figure S3. Kaplan-Meier plot of death-censored allograft survival for recipients with different numbers of APOL1 risk alleles | 4 |
| Figure S4. Recipient pAFR and creatinine in GOCAR and CTOT cohorts | 5 |
| Figure S5. Allele specific expression of APOL1 in single cell RNA sequencing data from PBMCs of two ESRD patients | 6 |


**Supplementary Tables**


| | |
|---|---|
| Table S1. Demographic and clinicopathologic characteristics of GOCAR and CTOT donors and recipients with genome-wide genotype data | 7-8 |
| Table S2. Genetic ancestry and self-reported ancestry of donors and recipients in CTOT | 9 |
| Table S3. APOL1 risk genotype in donor-recipient pairs of GOCAR and CTOT | 10 |
| Table S4. Summary of APOL1 risk alleles in GOCAR and CTOT cohorts stratified by recipients and donors with different genetic ancestries | 11 |
| Table S5. Association of APOL1 risk alleles with death-censored allograft loss in an additive manner in CTOT cohort | 12 |
| Table S6. Association of recipient APOL1 risk alleles with DCAL within the stratum of donors carrying APOL1 low-risk genotype in the GOCAR cohort. | 13 |
| Table S7. Association of recipient APOL1 risk alleles with DCAL within the stratum of donors carrying APOL1 low-risk genotype in the CTOT cohort. | 14 |
| Table S8. Association of APOL1 risk alleles with different TCMR outcomes in an additive way in CTOT data | 15 |
| Table S9. Association of APOL1 risk alleles with death-censored allograft loss independent of APOL1 SNP-wise mismatch in GOCAR and CTOT | 16 |
| Table S10. Enrichment in KEGG pathways of DEGs identified in immune cells in DICE data | 17 |
| Table S11. Enrichment in KEGG pathways of DEGs identified from two ESRD patients with single cell RNA sequencing data of PBMCs | 17 |
| Table S12. Enrichment in immune related pathways of DEGs identified from a subset of GOCAR recipients with PBMC RNA sequencing data | 17 |
| Table S13. Genes used to define cell types in the scRNAseq data analysis for two ESRD patients | 17 |



**References** 18

### Supplementary Figures

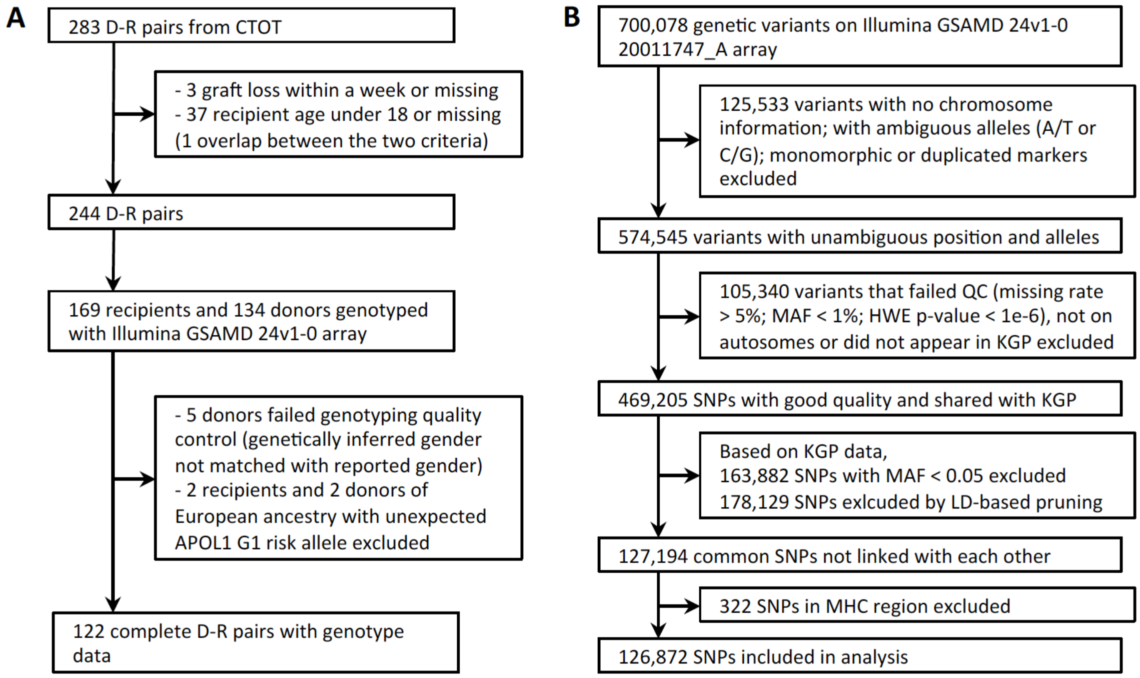


**Figure S1. Flowchart of genotyping and quality control for CTOT cohort. (A)** Participants; **(B)** Genetic variants. QC: quality control; MAF: minor allele frequency; HWE: Hardy-Weinberg equilibrium; KGP: 1000 Genomes Project; LD: linkage disequilibrium; MHC: major histocompatibility complex.

A

B

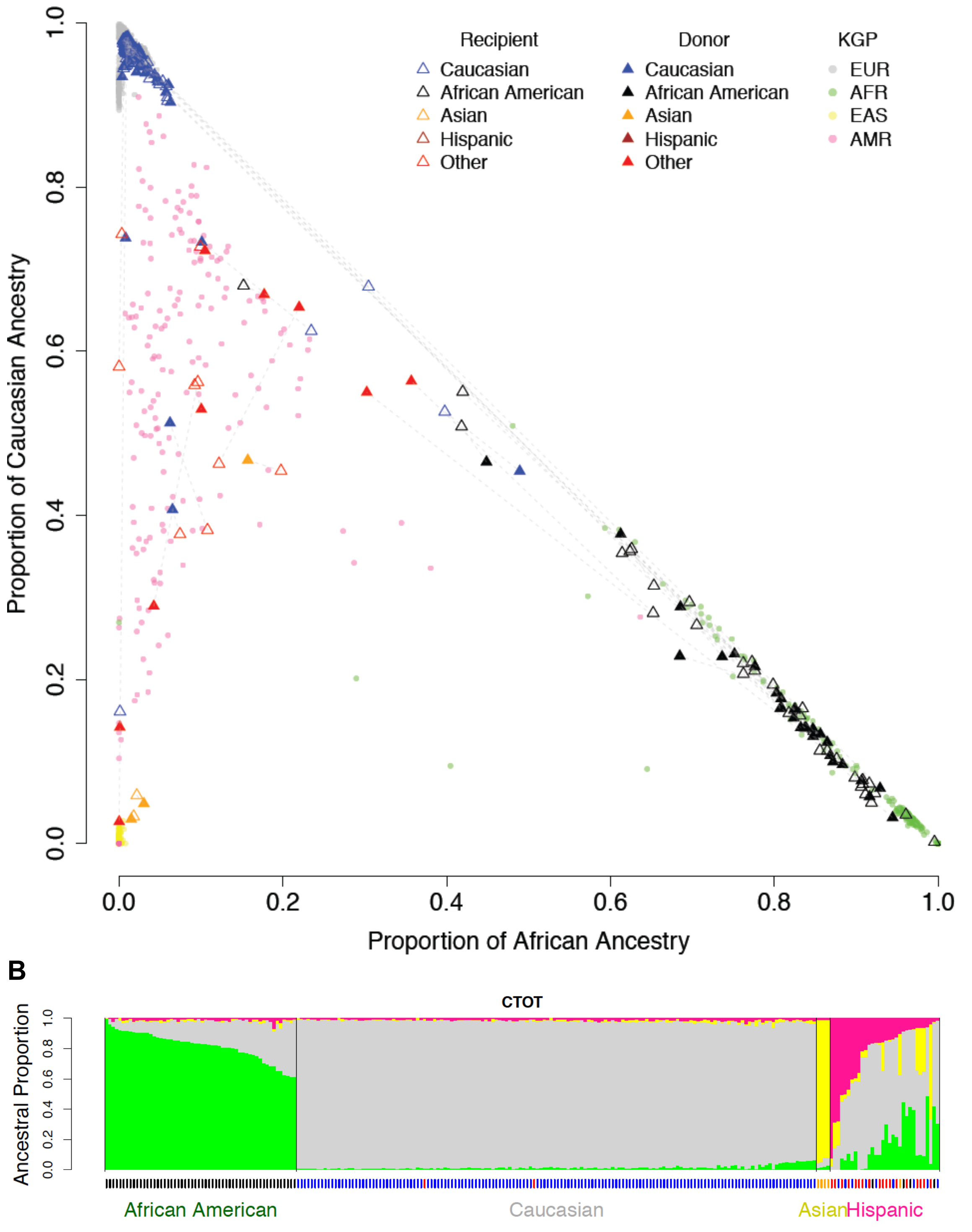


**Figure S2. Genetic ancestry of CTOT donors and recipients. (A)** Samples from 1000 Genomes Project (KGP) with different ethnicities anchor the location of continental-level ancestries on the space spanned by estimated proportions of African and Caucasian ancestries. CTOT donors (solid triangle) and recipients (empty triangle) are projected onto the same space and colored based on self-reported ancestry. Donor-recipient pairs are connected by dashed lines. KGP: 1000 Genomes Project; EUR: European; AFR: African; EAS: East Asian; AMR: American. **(B)** The ancestral composition of each individual in CTOT. Each vertical bar represents an individual. The length of colored segments within each bar indicates the estimated proportion of different genetic ancestries. The ticks under the bar plot indicates self-reported race with the same color code in legend of (A).

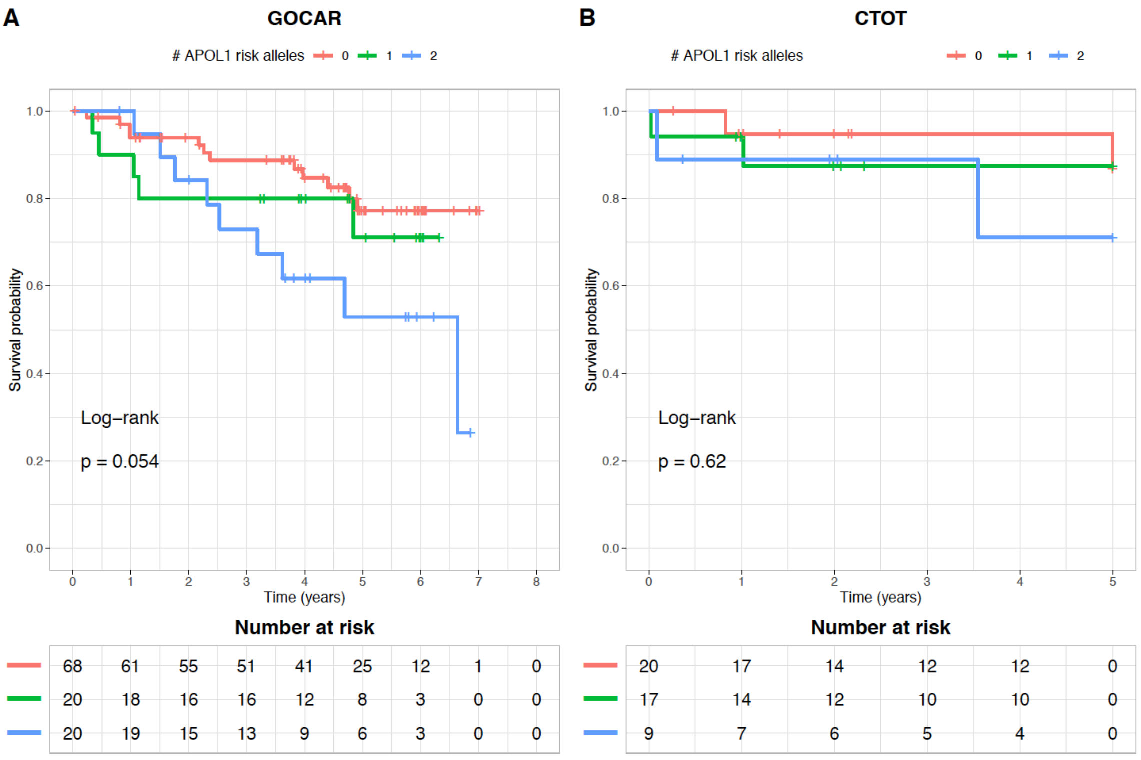


**Figure S3. Kaplan-Meier plot of death-censored allograft survival for recipients with different numbers of APOL1 risk alleles.** The subset of African American and Hispanic recipients was shown. (**A**) GOCAR; (**B**) CTOT.

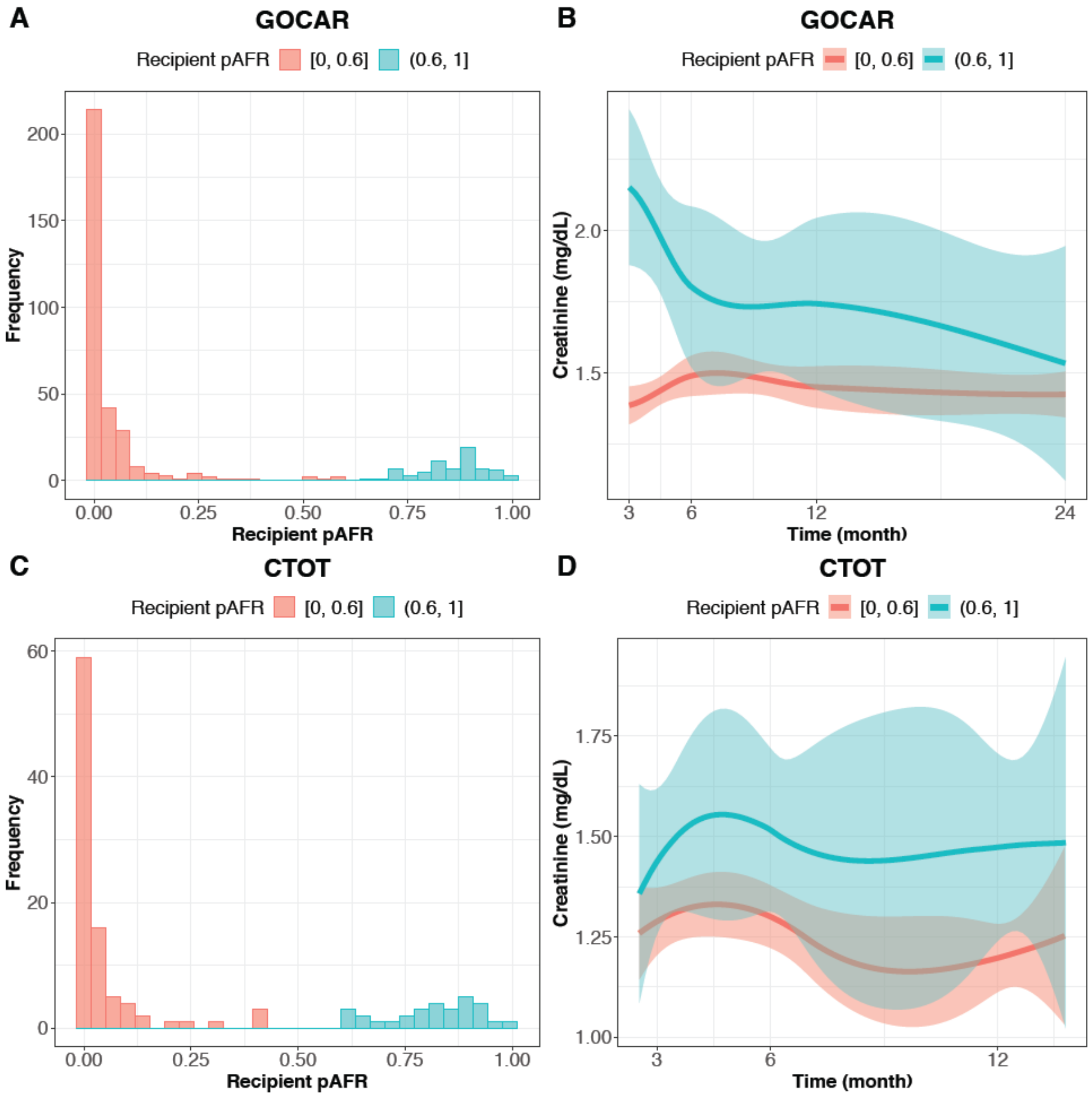


**Figure S4. Recipient pAFR and creatinine in GOCAR and CTOT cohorts.** (**A**) and (**C**) Histogram of the distribution of recipient pAFR values in GOCAR and CTOT cohorts. Recipients were categorized in 2 groups by their pAFR corresponding to non-African American and African American. (**B**) and (**D**) Smoothed curves with 95% confidence band for longitudinal creatinine levels grouped by recipient pAFR as shown in (A) and (C) for GOCAR and CTOT cohorts.

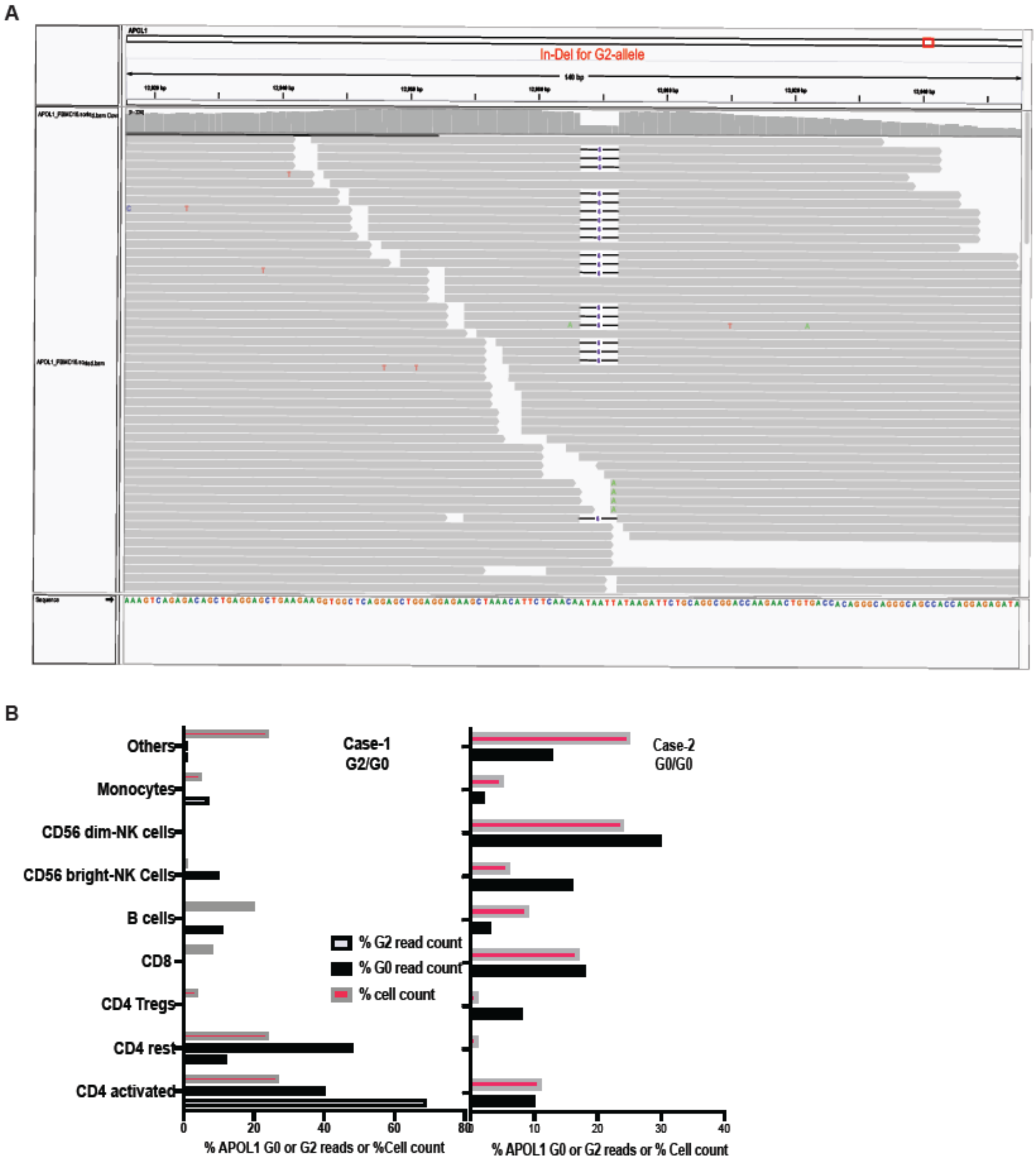


**Figure S5. Allele specific expression of APOL1 in single cell RNA sequencing data from PBMCs of two ESRD patients.** (**A**) Visualization of the expression of APOL1 G2 allele represented by the short reads (indicated by gray thick arrows) carrying the 6 bp micro-deletion (indicated by a short segment) in the patient with G2 allele using Integrative Genomics Viewer (IGV).[1] (**B**) The percentage of different types of immune cells and the percentage of short reads carrying G2 and G0 alleles in the two patients. Here, G0 refers to the APOL1 allele without G1/G2 risk variants.

**Supplementary Tables**

### Table S1. Demographic and clinicopathologic characteristics of GOCAR and CTOT donors and recipients with genome-wide genotype data.

| Variable | GOCAR D-R pairs with genotype (n = 385) [a] | CTOT D-R pairs with genotype (n = 122) [b] | p-value[c] |
|---|---|---|---|
| **Recipient** | | | |
| **Death censored graft loss** (years) | | | |
| mean ± SD; median (range) | 4.6 ± 1.7; 4.9 (0.04, 7.3) | 3.7 ± 1.8; 5.0 (0.02, 5.0) | **<0.001** |
| # events (%) | 50 (13.0%) | 6 (4.9%) | **0.01** |
| **TCMR >= borderline,** # events (%) | 126 (32.7%) | 15 (12.3%) | **<0.001** |
| **TCMR > borderline**, # events (%) | 36 (9.4%) | 1 (0.8%) | **<0.001** |
| **Recurrent TCMR >= borderline**, # events (%) | 59 (15.3%) | - | - |
| **Recurrent TCMR > borderline**, # events (%) | 25 (6.5%) | - | - |
| **Age** (years), mean ± SD; median (range) | 49.9 ± 13.5; 50 (18, 83) | 48.8 ± 13.6; 50 (18, 89) | 0.44 |
| **Gender**, male, n (%) | 257 (66.8%) | 74 (60.7%) | 0.23 |
| **Genetic ancestry**[d], n (%) | | | 0.30 |
| African American | 70 (18.2%) | 30 (24.6%) | |
| Asian | 13 (3.4%) | 2 (1.6%) | |
| Caucasian | 235 (61.0%) | 74 (60.7%) | |
| Hispanic | 67 (17.4%) | 16 (13.1%) | |
| **HLA mismatch score**[e], n (%) | 2.0 ± 1.0 | 3.3 ± 1.8 | **0.03**[†] |
| **Induction**, n (%) | | | 0.11 |
| No induction | 78 (20.3%) | 36 (29.5%) | |
| Non-depletional (IL2 antagonist) | 130 (33.8%) | 36 (29.5%) | |
| Depletional (Thymoglobulin or Campath) | 177 (46.0%) | 50 (41.0%) | |
| **# APOL1 risk alleles**[g], n (%) | | | **0.001** |
| 0 | 316 (82.1%) | 94 (77.0%) | |
| 1 | 20 (5.2%) | 17 (13.9%) | |
| 2 | 20 (5.2%) | 9 (7.4%) | |
| N/A | 29 (7.5%) | 2 (1.6%) | |

| | | | |
|---|---|---|---|
| **Donor** | | | |
| **Age** (years), mean ± SD; median (range) | 42.6 ± 14.7; 45 (3, 73) | 40.3 ± 12.3; 39 (6, 65) | 0.09 |
| **Gender**, male, n (%) | 196 (50.9%) | 49 (40.2%) | **0.05** |
| **Genetic ancestry**, n (%) | | | **0.003** |
| African American | 33 (8.6%) | 26 (21.3%) | |
| Asian | 7 (1.8%) | 2 (1.6%) | |
| Caucasian | 293 (76.1%) | 78 (63.9%) | |
| Hispanic | 52 (13.5%) | 16 (13.1%) | |
| **Donor type,** live donor, n (%) | 194 (50.4%) | 105 (86.8%) | **<0.001** |
| **# APOL1 risk alleles**[g], n (%) | | | **<0.001** |
| 0 | 355 (92.2%) | 99 (81.1%) | |
| 1 | 10 (2.6%) | 16 (13.1%) | |
| 2 | 6 (1.6%) | 5 (4.1%) | |
| N/A | 14 (3.6%) | 2 (1.6%) | |

[a]: Genome-wide genotype data is available for 385 donor-recipient (D-R) pairs from the parent GOCAR study after data processing and quality control detailed elsewhere.[2]

[b]: Genome-wide genotype data is available for 122 donor-recipient (D-R) pairs from the parent CTOT study after data processing and quality control (see Methods).

[c]: P-value was calculated from unpaired t-test for continuous variables and from Fisher's exact test for categorical variables unless otherwise specified. Bold p-value < 0.05.

[d]: Genetic ancestry was inferred from genome-wide genotype data and considered more accurate than self-reported race.[2]

[e]: HLA mismatch score was derived from 2-digit HLA allele typing. Following previous reports for GOCAR,[2-4] the raw mismatch score (scaling from 0 to 6) was categorized into: 0 (no mismatches), 1 (1-2 mismatches), 2 (3-4 mismatches), and 3 (5-6 mismatches); while for the CTOT cohort, the raw mismatch score (scaling from 0 to 6) was used. In subsequent statistical analyses, this variable was used as numeric covariate in regression models.

[f]: In order to calculate the p-value, the raw HLA mismatch score used in CTOT was hereby categorized in the same way as GOCAR so that the HLA mismatch scores originally defined on different scales in the two cohorts are comparable. The p-value was calculated by Fisher's exact test.

[g]: There are missing data in the APOL1 genotype of 29 recipients and 14 donors for GOCAR cohort and of 2 recipients and 2 donors for CTOT cohort (see supplementary Table S3 for details).

**Table S2. Genetic ancestry and self-reported ancestry of donors and recipients in CTOT.** Genetic ancestry is inferred from genome-wide genetic data for n = 122 D-R pairs.

**(a) Donor: Genetic ancestry versus self-reported race**

| | | Self-reported Race | | | | |
|---|---|---|---|---|---|---|
| | | African American | Asian | Caucasian | Hispanic | Other/ Unreported |
| Genetic Ancestry | African American | 26 | 0 | 0 | 0 | 0 |
| | Asian | 0 | 2 | 0 | 0 | 0 |
| | Caucasian | 0 | 0 | 76 | 0 | 2 |
| | Hispanic | 1 | 1 | 5 | 0 | 9 |

**(b) Recipient: Genetic ancestry versus self-reported race**

| | | Self-reported Race | | | | |
|---|---|---|---|---|---|---|
| | | African American | Asian | Caucasian | Hispanic | Other/ Unreported |
| Genetic Ancestry | African American | 30 | 0 | 0 | 0 | 0 |
| | Asian | 0 | 2 | 0 | 0 | 0 |
| | Caucasian | 0 | 0 | 74 | 0 | 0 |
| | Hispanic | 3 | 0 | 4 | 0 | 9 |

**(c) Donor and recipient genetic ancestry**

| | | Recipient Genetic Ancestry | | | | |
|---|---|---|---|---|---|---|
| | | African American | Asian | Caucasian | Hispanic | Total |
| Donor Genetic Ancestry | African American | 24 | 0 | 1 | 1 | 26 |
| | Asian | 0 | 2 | 0 | 0 | 2 |
| | Caucasian | 4 | 0 | 72 | 2 | 78 |
| | Hispanic | 2 | 0 | 1 | 13 | 16 |
| | Total | 30 | 2 | 74 | 16 | 122 |

**Table S3. APOL1 risk genotype in donor-recipient pairs of GOCAR and CTOT.**

**(A) GOCAR (n = 385 D-R pairs)**

| | | Recipient APOL1 risk genotype | | | | |
|---|---|---|---|---|---|---|
| | | G0/G0 | G0/G1 or G0/G2 | G1/G1, G1/G2, or G2/G2 | N/A | Total |
| Donor APOL1 risk genotype | G0/G0 | 305 | 17 | 14 | 19 | 355 |
| | G0/G1 or G0/G2 | 6 | 0 | 1 | 3 | 10 |
| | G1/G1, G1/G2, or G2/G2 | 0 | 2 | 2 | 2 | 6 |
| | N/A | 5 | 1 | 3 | 5 | 14 |
| | Total | 316 | 20 | 20 | 29 | 385 |

**(B) CTOT (n = 122 D-R pairs)**

| | | Recipient APOL1 risk genotype | | | | |
|---|---|---|---|---|---|---|
| | | G0/G0 | G0/G1 or G0/G2 | G1/G1, G1/G2, or G2/G2 | N/A | Total |
| Donor APOL1 risk genotype | G0/G0 | 87 | 7 | 5 | 0 | 99 |
| | G0/G1 or G0/G2 | 5 | 8 | 3 | 0 | 16 |
| | G1/G1, G1/G2, or G2/G2 | 2 | 2 | 1 | 0 | 5 |
| | N/A | 0 | 0 | 0 | 2 | 2 |
| | Total | 94 | 17 | 9 | 2 | 122 |

**Table S4. Summary of APOL1 risk alleles in GOCAR and CTOT cohorts stratified by recipients and donors with different genetic ancestries.**

| Genetic Ancestry | Recipient #APOL1 risk alleles | | | | Total | Donor #APOL1 risk alleles | | | | Total |
|---|---|---|---|---|---|---|---|---|---|---|
| | 0 | 1 | 2 | N/A | | 0 | 1 | 2 | N/A | |
| **GOCAR** (n = 385 D-R pairs) | | | | | | | | | | |
| African American | 12 | 16 | 15 | 27 | 70 | 9 | 7 | 6 | 11 | 33 |
| Asian | 13 | 0 | 0 | 0 | 13 | 7 | 0 | 0 | 0 | 7 |
| Caucasian | 235 | 0 | 0 | 0 | 235 | 293 | 0 | 0 | 0 | 293 |
| Hispanic | 56 | 4 | 5 | 2 | 67 | 46 | 3 | 0 | 3 | 52 |
| All | 316 | 20 | 20 | 29 | 385 | 355 | 10 | 6 | 14 | 385 |
| **CTOT** (n = 122 D-R pairs) | | | | | | | | | | |
| African American | 8 | 14 | 8 | 0 | 30 | 6 | 15 | 5 | 0 | 26 |
| Asian | 2 | 0 | 0 | 0 | 2 | 2 | 0 | 0 | 0 | 2 |
| Caucasian | 72 | 0 | 0 | 2 | 74 | 76 | 0 | 0 | 2 | 78 |
| Hispanic | 12 | 3 | 1 | 0 | 15 | 15 | 1 | 0 | 0 | 16 |
| All | 94 | 17 | 9 | 2 | 122 | 99 | 16 | 5 | 2 | 122 |

**Table S5. Association of APOL1 risk alleles with death-censored allograft loss in an additive manner in CTOT cohort.**

| Variable[a] | HR | 95% CI | p-value[d] |
|---|---|---|---|
| ***CTOT: recipients all ancestries[b] (n = 117[c]; 6 [5.1%] graft loss events)*** | | | |
| **# APOL1 risk alleles** | 2.73 | (1.04, 7.20) | **0.04** |
| **Donor type** (ref: LD) DD | 3.85 | (0.71, 20.8) | 0.12 |
| **HLA mismatch score** | 1.16 | (0.73, 1.83) | 0.52 |
| ***CTOT: recipients of African American and Hispanic (n = 46[c]; 6 [13.0%] graft loss events)*** | | | |
| **# APOL1 risk alleles** | 1.32 | (0.46, 3.81) | 0.60 |
| **Donor type** (ref: LD) DD | 3.96 | (0.70, 22.4) | 0.12 |
| **HLA mismatch score** | 1.08 | (0.64, 1.84) | 0.77 |

[a]: In the multivariable Cox regression model, donor type and HLA mismatch score were forced into model, while other covariates adjusted in GOCAR data, including recipient ancestry and induction, that were not significant in multivariable analysis were not included in order to increase statistical power for the CTOT cohort with limited sample size.

[b]: The “Asian” category was excluded due to limited sample size which led to instable model fitting.

[c]: Sample size was reduced due to missing data in donor APOL1 risk alleles.

[d]: Bold p-value < 0.05.

**Table S6. Association of recipient APOL1 risk alleles with death-censored allograft loss using multivariable Cox regression, within the stratum of donors carrying APOL1 low-risk genotype in the GOCAR cohort.**

| Variable | HR | 95% CI | p-value[d] |
|---|---|---|---|
| ***GOCAR: recipients of all ancestries[a], within the stratum of donors carrying APOL1 low-risk genotype[b] (n = 330 [c]; 41 [12.4%] graft loss events)*** | | | |
| **# APOL1 risk alleles** | 1.93 | (1.06, 3.49) | **0.03** |
| **Recipient genetic ancestry** (ref: Caucasian) | | | |
| **African American** | 1.02 | (0.29, 3.57) | 0.98 |
| **Hispanic** | 2.70 | (1.23, 5.95) | **0.01** |
| **Induction** (ref: No) | | | |
| **Non-depletional** | 2.84 | (0.92, 8.79) | 0.07 |
| **Depletional** | 3.56 | (1.14, 11.1) | **0.03** |
| **Donor type** (ref: LD) DD | 2.62 | (1.24, 5.51) | **0.01** |
| **HLA mismatch score** | 1.20 | (0.81, 1.77) | 0.36 |
| ***GOCAR: recipients of African American and Hispanic, within the stratum of donors carrying APOL1 low-risk genotype[b] (n = 97 [c]; 23 [23.7%] graft loss events)*** | | | |
| **# APOL1 risk alleles** | 2.11 | (1.14, 3.88) | **0.02** |
| **Recipient genetic ancestry** (ref: AA) Hispanic | 2.98 | (0.90, 9.82) | 0.07 |
| **Induction** (ref: No) | | | |
| **Non-depletional** | 5.46 | (0.62, 47.7) | 0.13 |
| **Depletional** | 4.93 | (0.60, 40.7) | 0.14 |
| **Donor type** (ref: LD) DD | 2.90 | (0.88, 9.55) | 0.08 |
| **HLA mismatch score** | 1.67 | (0.85, 3.30) | 0.14 |

[a]: The “Asian” category was excluded due to limited sample size which led to instable model fitting.

[b]: Donor APOL1 high-risk genotype is defined as 2 copies of G1/G2 alleles and low-risk genotype as 0 or 1 G1/G2 allele.

[c]: Sample size was reduced due to missing data in APOL1 risk alleles.

[d]: Bold p-value < 0.05.

**Table S7. Association of recipient APOL1 risk alleles with death-censored allograft loss using multivariable Cox regression, within the stratum of donors carrying APOL1 low-risk genotype in the CTOT cohort.**

| Variable[a] | HR | 95% CI | p-value[e] |
|---|---|---|---|
| ***CTOT: recipients all ancestries[b], within the stratum of donors carrying APOL1 low risk genotype[c] (n = 112[d]; 6 [5.4%] graft loss events)*** | | | |
| **# APOL1 risk alleles** | 2.84 | (1.05, 7.70) | **0.04** |
| **Donor type** (ref: LD) DD | 3.61 | (0.67, 19.4) | 0.13 |
| **HLA mismatch score** | 1.13 | (0.71, 1.79) | 0.62 |
| ***CTOT: recipients of African American and Hispanic, within the stratum of donors carrying APOL1 low risk genotype[c] (n = 41[d]; 6 [14.6%] graft loss events)*** | | | |
| **# APOL1 risk alleles** | 1.32 | (0.45, 3.91) | 0.62 |
| **Donor type** (ref: LD) DD | 3.76 | (0.67, 21.1) | 0.13 |
| **HLA mismatch score** | 1.04 | (0.60, 1.80) | 0.88 |

[a]: In the multivariable Cox regression model, donor type and HLA mismatch score were forced into model, while other covariates adjusted in GOCAR data, including recipient ancestry and induction, that were not significant in multivariable analysis were not included in order to increase statistical power for the CTOT cohort with limited sample size.

[b]: The "Asian" category was excluded due to limited sample size which led to instable model fitting.

[c]: Donor APOL1 high-risk genotype is defined as 2 copies of G1/G2 alleles and low-risk genotype as 0 or 1 G1/G2 allele.

[d]: Sample size was reduced due to missing data in donor APOL1 risk alleles.

[e]: Bold p-value < 0.05.

**Table S8. Association of recipient APOL1 risk alleles with different TCMR outcomes in the CTOT cohort.**

| TCMR outcome | $n_{control}$ [a,b] | $n_{case}$ [a] | OR | 95% CI | p-value[d] |
|---|---|---|---|---|---|
| ***CTOT: recipient of all ancestries*** | | | | | |
| **TCMR >= borderline** (Univariate) | 106 | 14 | 2.32 | (1.06, 4.87) | **0.03** |
| **TCMR >= borderline** (Multivariable[c]) | 100 | 14 | 2.32 | (1.02, 5.16) | **0.04** |
| ***CTOT: recipients of African American and Hispanic*** | | | | | |
| **TCMR >= borderline** (Univariate) | 39 | 7 | 2.95 | (1.01, 10.3) | 0.06 |
| **TCMR >= borderline** (Multivariable[c]) | 34 | 7 | 3.39 | (1.07, 13.6) | **0.05** |

[a]: Sample size was reduced due to missing data in APOL1 risk alleles for univariate analysis and due to missing data in APOL1 risk alleles and HLA mismatch score for multivariable analysis.
[b]: Controls (no TCMR) were defined as patients with either (a) no TCMR or borderline TCMR on obtained biopsies at anytime, or (b) no reported biopsies during follow up.
[c]: In the multivariable logistic regression model, we focused on the stratum of donors with APOL1 low-risk genotype carrying 0 or 1 G1/G2 allele, because the model fitting would not have converged if the donor APOL1 risk genotype had been included as a covariate due to the limited number (n = 5) of donors with APOL1 high-risk genotype carrying 2 G1/G2 alleles. HLA mismatch score were forced into model, while other covariates adjusted in GOCAR data, including recipient ancestry, induction, and donor type that were not significant in multivariable analysis were not included in order to increase statistical power for the CTOT cohort with limited sample size.
[d]: Bold p-value < 0.05.

**Table S9. Association of APOL1 risk alleles with death-censored allograft loss independent of APOL1 SNP-wise mismatch in GOCAR and CTOT.**

| Variable[a] | HR | 95% CI | p-value[d] |
|---|---|---|---|
| ***GOCAR: recipients of all ancestries[b] (n = 343[c]; 44 [12.8%] graft loss events)*** | | | |
| **# APOL1 risk alleles** | 2.24 | (1.30, 3.86) | **0.004** |
| **APOL1 SNP-wise mismatch** | 0.75 | (0.40, 1.44) | 0.39 |
| ***GOCAR: recipients of African American and Hispanic (n = 108[c]; 26 [24.1%] graft loss events)*** | | | |
| **# APOL1 risk alleles** | 2.46 | (1.38, 4.40) | **0.002** |
| **APOL1 SNP-wise mismatch** | 0.75 | (0.32, 1.76) | 0.51 |
| ***CTOT: recipients of all ancestries (n = 117[c]; 6 [5.1%] graft loss events)*** | | | |
| **# APOL1 risk alleles** | 2.56 | (0.98, 6.73) | 0.06 |
| **APOL1 SNP-wise mismatch** | 1.39 | (0.73, 2.66) | 0.31 |
| ***CTOT: recipients of African American and Hispanic (n = 46[c]; 6 [13.0%] graft loss events)*** | | | |
| **# APOL1 risk alleles** | 1.10 | (0.35, 3.41) | 0.87 |
| **APOL1 SNP-wise mismatch** | 1.62 | (0.78, 3.35) | 0.19 |

[a]: In the Cox regression model for GOCAR, covariates include recipient ancestry, number of APOL1 risk alleles, and APOL1 SNP-wise mismatch, induction, donor type, and HLA mismatch score. In the multivariable Cox regression model for CTOT, donor type and HLA mismatch score were forced into model, while other covariates adjusted in GOCAR data, including recipient ancestry and induction, that were not significant in multivariable analysis were not included in order to increase statistical power for the CTOT cohort with limited sample size. For concise presentation, only recipient number of APOL1 risk alleles and APOL1 SNP-wise mismatch were shown in the table.

[b]: The "Asian" category was excluded due to limited sample size which led to instable model fitting.

[c]: Sample size was reduced due to missing data in APOL1 risk alleles.

[d]: Bold p-value < 0.05.

**Table S10. Enrichment in KEGG pathways of DEGs identified in immune cells in DICE data.**
(Supplementary_Tables_S10_to_S12.xlsx)

**Table S11. Enrichment in KEGG pathways of DEGs identified from two ESRD patients with single cell RNA sequencing data of PBMCs.**
(Supplementary_Tables_S10_to_S12.xlsx)

**Table S12. Enrichment in immune related pathways of DEGs identified from a subset of GOCAR recipients with PBMC RNA sequencing data.**
(Supplementary_Tables_S10_to_S12.xlsx)

**Table S13. Genes used to define cell types in the scRNAseq data analysis for two ESRD patients.**

| Cell type | Genes used to define cell type |
|---|---|
| Activated $CD4^+$ T cell | CD3D, CD3E, LTB, CD4, TNF, STAT1, MAF |
| Activated $CD8^+$ T cell | CD3D, CD3E, CD8A, CD8B, GZMB, GNLY, PRF1 |
| $CD56^{dim}$ NK cell | GNLY, NKG7, PRF1, GZMB, GZMH, FGFBP2 |
| Monocyte | CD14, LYZ, S100A8, S100A9 |
| B Cell | CD19, CD79A, CD74, MS4A1 |
